# The impact of vaccinating adolescents and children on COVID-19 disease outcomes

**DOI:** 10.1101/2021.10.21.21265318

**Authors:** Kylie E. C. Ainslie, Jantien Backer, Pieter de Boer, Albert Jan van Hoek, Don Klinkenberg, Hester Korthals Altes, Ka Yin Leung, Hester de Melker, Fuminari Miura, Jacco Wallinga

**Affiliations:** Centre for Infectious Disease Control, National Institute for Public Health and the Environment, Bilthoven, the Netherlands; School of Public Health, Imperial College London, London, United Kingdom; MRC Centre for Global Infectious Disease Analysis and Abdul Latif Jameel Institute for Disease and Emergency Analytics, Imperial College London, London, United Kingdom; Center for Marine Environmental Studies (CMES), Ehime University, Ehime, Japan; Department of Biomedical Data Sciences, Leiden University Medical Center, Leiden, the Netherlands

## Abstract

**Introduction:** Despite the high COVID-19 vaccination coverage among adults, there is concern over a peak in SARS-CoV-2 infections in the coming months. To help ensure that healthcare systems are not overwhelmed in the event of a new wave of SARS-CoV-2 infections, many countries have extended vaccination to adolescents (those aged 12-17 years) and may consider further extending to children aged 5-11 years. However, there is considerable debate about whether or not to vaccinate healthy adolescents and children against SARS-CoV-2 because, while vaccination of children and adolescents may limit transmission from these groups to other, more vulnerable groups, adolescents and children themselves have limited risk of severe disease if infected and may experience adverse events from vaccination. To quantify the benefits of extending COVID-19 vaccination beyond adults we compare daily cases, hospital admissions, and intensive care (IC) admissions for vaccination in adults only, those 12 years and above, and those 5 years and above.

**Methods and Findings:** We developed a deterministic, age-structured susceptible-exposed-infectious-recovered (SEIR) model to simulate disease outcomes (e.g., cases, hospital admissions, IC admissions) under different vaccination scenarios. The model is partitioned into 10-year age bands (0-9, 10-19, …, 70-79, 80+) and accounts for differences in susceptibility and infectiousness by age group, seasonality in transmission rate, modes of vaccine protection (e.g., infection, transmission), and vaccine characteristics (e.g., vaccine effectiveness). Model parameters are estimated by fitting the model piecewise to daily cases from the Dutch notification database Osiris from 01 January 2020 to 22 June 2021. Forward simulations are performed from 22 June 2021 to 31 March 2022. We performed sensitivity analyses in which vaccine-induced immunity waned.

We found that upon relaxation of all non-pharmaceutical control measures a large wave occurred regardless of vaccination strategy. We found overall reductions of 5.7% (4.4%, 6.9%) of cases, 2.0% (0.7%, 3.2%) of hospital admissions, and 1.7% (0.6%, 2.8%) of IC admissions when those 12 years and above were vaccinated compared to vaccinating only adults. When those 5 years and above were vaccinated we observed reductions of 8.7% (7.5%, 9.9%) of cases, 3.2% (2.0%, 4.5%) of hospital admissions, and 2.4% (1.2%, 3.5%) of IC admissions compared to vaccination in adults only. Benefits of extending vaccination were larger within the age groups included in the vaccination program extension than in other age groups. The benefits of vaccinating adolescents and children were smaller if vaccine protection against infection, hospitalization, and transmission (once infected) wanes.

**Discussion:** Our results highlight the benefits of extending COVID-19 vaccination programs beyond adults to reduce infections and severe outcomes in adolescents and children and in the wider population. A reduction of infections in school-aged children/adolescents may have the added benefit of reducing the need for school closures during a new wave. Additional control measures may be required in future to prevent a large wave despite vaccination program extensions. While the results presented here are based on population characteristics and the COVID-19 vaccination program in The Netherlands, they may provide valuable insights for other countries who are considering COVID-19 vaccination program extensions.

## Introduction

Despite the high COVID-19 vaccination coverage among adults in several developed countries, there is concern over future waves of infections. As COVID-19 vaccines have been approved for adolescents [1-3] and are being considered for approval in children [4], one of the main policy decisions under consideration is extending vaccination beyond adults. Increased transmission may be due to seasonality of transmission [5-7], possible waning of vaccine protection [8-10], and reduced vaccine effectiveness against variants [11-14]. The high transmissibility of the Delta variant, now the dominant strain in Europe and the United States [15], makes reaching herd immunity by achieving a high vaccination coverage in adults unlikely. Therefore, many countries are shifting policy towards managing incidence of severe disease. To that end, many developed countries have extended COVID-19 vaccination eligibility to adolescents (12-17 year olds), particularly those with comorbidities [16-21]. However, there is considerable debate about whether to vaccinate healthy adolescents.

Healthy adolescents can be infected by and transmit SARS-CoV-2 [22], but are much less likely to experience severe disease [23-25] and die [26, 27] following infection with SARS-CoV-2 compared to adults. Additionally, some vaccinated adolescents may experience adverse events, such as pericarditis and myocarditis (heart inflammation), following vaccination with a COVID-19 vaccine [28, 29]. Despite their reduced risks of severe outcomes, adolescents may experience symptoms lasting months after infection (‘long COVID’) [30-32]. In the Netherlands, the disease burden of COVID-19 among adolescents (excluding ‘long COVID’) has been shown to be similar to seasonal influenza [33], while in the United States, adolescent hospitalization rates due to COVID-19 were 2.5–3.0 times higher than those from the three recent influenza seasons [34]. Therefore, a direct benefit of vaccinating adolescents will be to reduce incidence of severe disease and long COVID, as well as infections, in this age group. Additional benefits of vaccinating adolescents is reducing the chance of school closures during a future wave if a high proportion of students are vaccinated [21]. School closures have been used throughout the pandemic to limit spread and have been shown to be very disruptive to adolescents [35-37] and their parents, resulting in high economic costs [37]. Therefore, vaccination of this group may have higher direct benefits in terms of access to education and mental well-being [21].

Another objective for vaccinating adolescents is to reduce transmission from this group to other, more vulnerable, groups. Adolescents make a high number of daily contacts [38-40] and have lower adherence to non-pharmaceutical interventions [41]. Therefore, this group is likely to be a large contributor to transmission in the event of large outbreaks, as was seen in late June and early July in the Netherlands [42]. In the United Kingdom, an increase in infections in adolescents and young adults preceded the second and third waves, where infections later spread to older age groups [43-46]. While data on vaccine effectiveness against transmission in adolescents is currently lacking, studies have shown that household members of vaccinated adults were at reduced risk of infection with SARS-CoV-2 [47-49]. If vaccination of adolescents provides similar protection against transmission to household members, then vaccination may be a useful tool to reduce transmission from adolescents to other groups.

As with adolescents, children are at a reduced risk of severe disease after infection with SARS-CoV-2 [23-25]. However, vaccination of children may reduce transmission of SARS-CoV-2 in the wider population, as has been shown in the context of influenza [50]. Prior COVID-19 modelling work has shown that high incidence of SARS-CoV-2 infections may be expected if children are not vaccinated and that this will contribute to infections in unprotected adults [51, 52]. Therefore, fewer non-pharmaceutical interventions, such as school closures, would be required to control infection rates if children were also included in the vaccination program [52].

Little evidence has been published thus far about the effects of vaccinating adolescents or further extending COVID-19 vaccination programs to include children. In this work, we quantify the impacts of extending COVID-19 vaccination beyond adults using a deterministic, age-structured susceptible-exposed-infectious-recovered (SEIR) model. Our approach was previously used to inform vaccination policy regarding the vaccination of 12-17 year olds in The Netherlands [16, 33] before the emergence of the Delta variant. We therefore use the Netherlands as our motivating example. Here, we present updated analyses using model parameters consistent with the Delta variant. We quantified the impacts of vaccinating adolescents (12-17 years olds) and adolescents and children (5-17 year olds) on disease outcomes (e.g., cases, hospital admissions, and IC admissions). We compare disease outcomes in extended vaccination scenarios to disease outcomes when only adults are vaccinated. We performed sensitivity analyses to determine the impact of extending vaccination to adolescents in the present of waning vaccine-induced immunity.

## Methods

### Model Description

We developed a deterministic age-structured compartmental susceptible-exposed-infectious-recovered (SEIR) model extended to include states for severe disease outcomes and vaccination status. The population is partitioned into 10-year age groups (0-9, 10-19, …, 70-79, 80+). Within each age group we further partition the population into those who are unvaccinated, vaccinated with 1 dose, or vaccinated with 2 doses and then finally into disease states: susceptible (S), infected but not yet infectious (E), infectious (I), hospitalized (H), in intensive care (IC), return to the hospital ward after intensive care (H_IC_), recovered (R), and dead (D) (Figure S1). We assume that individuals who recover from infection cannot be re-infected. When a person is vaccinated, they first enter a hold state where they are vaccinated, but not yet (fully) protected (S_hold1d_ or S_hold2d_). After a delay period, they enter the vaccinated and protected state for the dose they have received (S_v1d_ or S_v2d_). We assume that vaccination only affects susceptible individuals.

The model is designed to incorporate a single vaccine product with a 2-dose regimen that 1) reduces susceptibility to infection, 2) reduces risk of hospitalization if a vaccinated individual is infected, and 3) reduces risk of infecting others (transmission) if a vaccinated person is infected. The vaccine is assumed to provide “leaky” protection, which means that the vaccine reduces the probability of transitioning between states for each vaccinated person. For example, a vaccine with 70% effectiveness against infection after the first dose would reduce the probability of transitioning from S_v1d_ to E_v1d_ by 70% compared to someone who is unvaccinated (moving from the S compartment to the E compartment). However, since there is more than one vaccine product currently licensed for use against COVID-19, we incorporate different vaccine products by taking the daily weighted average of the number of people with each vaccine product (and dose), the corresponding delay to protection of each vaccine product, and the vaccine effectiveness against each outcome (Table S1, see Supplemental Material for more details). Rate of vaccination for each day, vaccine product, and dose for each age group is a model input (see Supplemental Material for details).

The model uses different contact matrices from the Pienter Corona Study [38, 53, 54] estimated throughout 2020 and 2021 to approximate contact patterns under different levels of non-pharmaceutical interventions within and between age groups. These contact matrices are converted to transmission matrices by multiplying rows and columns by estimates of the relative susceptibility and infectiousness of each age group compared to the 0-9 years age group (Table S4).

### Seasonality

Seasonal cycles are a well-known feature of many respiratory viral infections, such as influenza and respiratory syncytial virus (RSV) [55-58]. Studies have shown that meteorological factors, such as temperature, specific humidity, and ultra violet (UV) radiation affect transmission of SARS-Cov-2 in temperate climates; however the amplitude of seasonal variability varies [5-7]. To account for the seasonal pattern of SARS-CoV-2 whereby, transmission is lower in summer and higher in winter, we define the transmission rate at time t, *β(t)*, as a sinusoidal function of seasonality [6] (see Supplemental Material for details)

### Model fit

The baseline (non-seasonal) transmission rate *β*_0_ and initial conditions for forward simulations are estimated by fitting the model to daily cases from the national notification database Osiris from 01 January 2020 to 22 June 2021, when vaccination in 12-17 year olds began in The Netherlands (Figure S2). The model is fitted to data piecewise to correspond with the correct contact patterns associated with different non-pharmaceutical interventions within each time window (Table S2). We directly estimate effective reproduction number (R_t_) within each time window using maximum likelihood estimation. We assume daily cases follow a negative binomial distribution with mean μ and overdispersion parameter *φ*. Estimates of R_t_ are converted to transmission rate by: 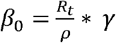, where γ is the inverse of the infectious period and ρ is the dominant eigen value of the product of the diagonal matrix of the number of susceptibles and the transmission matrix.

### Simulations

Forward simulations are performed from 22 June 2021 to 31 March 2022 with the initial conditions (i.e., the number of people in each compartment when the simulation begins) based on the last day of the model fitting. We assume the simulations begin with the same baseline transmission rate that was estimated from the last fitted time window (05 June to 22 June) and with contact patterns estimated during June 2021. We assume all non-pharmaceutical control measures are relaxed on 1 November 2021 for the remainder of the simulation period. Therefore, we assume the contact patterns change to those estimated prior to the COVID-19 pandemic in April 2017. We replicate our original analysis used to inform vaccination policy in the Netherlands when the Alpha variant was still the dominant variant (see Supplemental Material for details). At the time of writing, the Delta variant has become the dominant strain in Europe and the United States, so we update our analysis and perform simulations in which the baseline transmission rate in the absence of other non-pharmaceutical interventions is consistent with the basic reproduction number of the Delta variant (R_0_=4.6, *β*_0_ 0.*00078*). To incorporate uncertainty in the transmission rate, R_0_ is drawn from a normal distribution with mean 4.6 and standard deviation 0.0097 (corresponding to the estimated standard deviation of the effective reproduction number from the last time window during model fit), and converted to *β*_0_. Values of vaccine effectiveness against infection, hospitalization and transmission with the Delta variant were also updated (Table S1).

It is still unknown whether, and how much, vaccine effectiveness for vaccines against SARS-CoV-2 wanes over time. We look at two extremes: first, we assume no waning of vaccine-induced immunity; second, we include waning vaccine effectiveness as a logistic function, parameterized so that after 6 months vaccine-induced protection is reduced by 50% and by 99.6% after 1 year (after which the amount of waning is set to 100%). All vaccine types wane at the same rate for all outcomes. We do not assume waning of immunity from infection.

The objective of this specific model is to capture the dynamic aspects of vaccine allocation when comparing alternative vaccination strategies in children and adolescents. We assume vaccination of children and adolescents does not impact the continued vaccination of other groups. In the scenario in which 12–17 year olds are vaccinated (12+), we assume they receive the vaccines made by Pfizer/BioNTech and Moderna beginning 22 June 2021 and reach an overall coverage of 75% by 23 August 2021 as per the Dutch vaccination distribution schedule (Figure S3). When we extend vaccination to 5-11 year olds (5+), we assume a final vaccination coverage of 75%, a vaccine allocation of 50,000 doses per day, starting on 24 October 2021, and that they only receive the vaccine made by Pfizer/BioNTech.

We perform 200 simulations for each vaccination scenario. For each simulation we sample from the posterior distribution of the contact matrices to incorporate uncertainty regarding contact patterns [59]. We simulate new cases (assuming a case ascertainment rate of 33% per day), hospital admissions, and IC admissions for each vaccination strategy. We calculate the cumulative sum of each outcome for the entire simulation period. We also calculate the absolute and percent differences in each disease outcome for the different vaccination scenarios (12+ and 5+) compared to no vaccination in adults only (18+). Due to the stratification of the model population in 10-year age bands, we cannot separate 12–17 year olds from the 10–19 year age group or 5-11 year olds from the 0-9 and 10-19 age groups; therefore, we report the effects of the different vaccination strategies separately on the entire population, on the 0-9 age group, the 10-19 age group, and all individuals aged 20 and above. It is important to note the effect of vaccinating 12–17 year olds on the entire 10–19 year age group, includes the direct effects of vaccination on 12–17 year olds and the indirect effects on the 10–11 and 18– 19 year olds. Similarly, the effect of vaccinating 5-11 year olds on the entire 0-9 year age group includes the direct effect of vaccination on 5-9 year olds and the indirect effect on the 0-4 year olds.

The model is coded in R 4.1.0 [60] as a system of ordinary differential equations that are numerically solved using the lsoda function in the deSolve package [61]. For the full set of model equations see Supplemental Material. A full list of model input parameters are shown in Table S2 and Table S3. Code is available in the R package vacamole at https://github.com/kylieainslie/vacamole.

## Results

Our first observation is that after non-pharmaceutical control measures were released on 1 November 2021 there was a large wave of cases in all age groups regardless of vaccination strategy (Figure 1). When looking at the dynamics of disease outcomes by age group we saw that, in the absence of waning, 10-19 year olds were among the age groups with the highest incidence of cases when not included in the vaccination program (Figure 2s, 18+ panel, red line). However, when adolescents were included in the vaccination program, their incidence of cases decreased below other adult age groups (Figure 2, 5+ and 12+ panels, red line).

**Figure 1.**
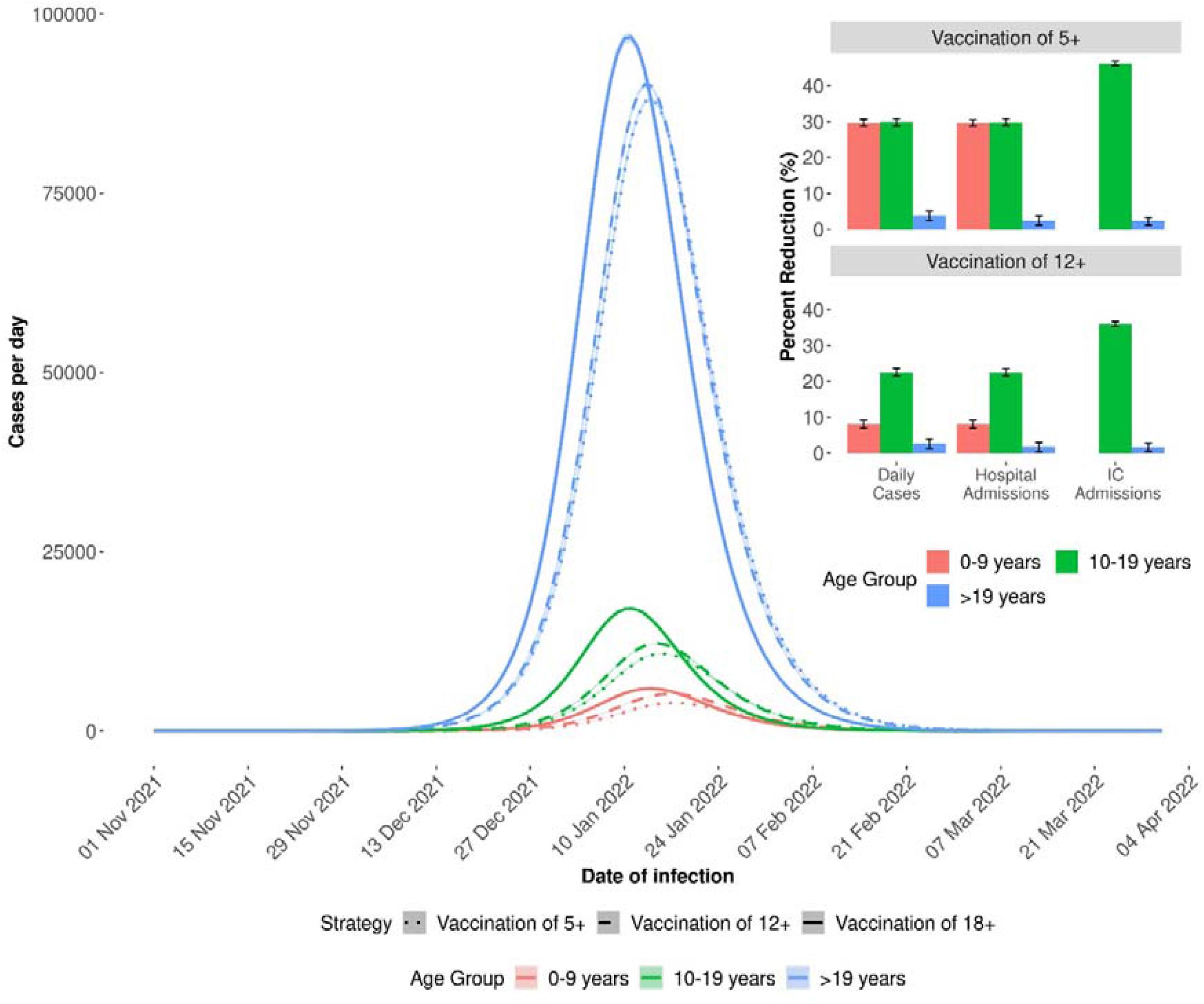
Daily cases for each vaccination scenario (5+, 12+, 18+) by age group (main) and percent reductions in disease outcomes in 5+ and 12+ vaccination scenarios compared to 18+ scenario (inset). Simulations were run from 22 June 2021 until 31 March 2022 with constant vaccine effectiveness. Control measures are relaxed on 1 November 2021.

**Figure 2.**
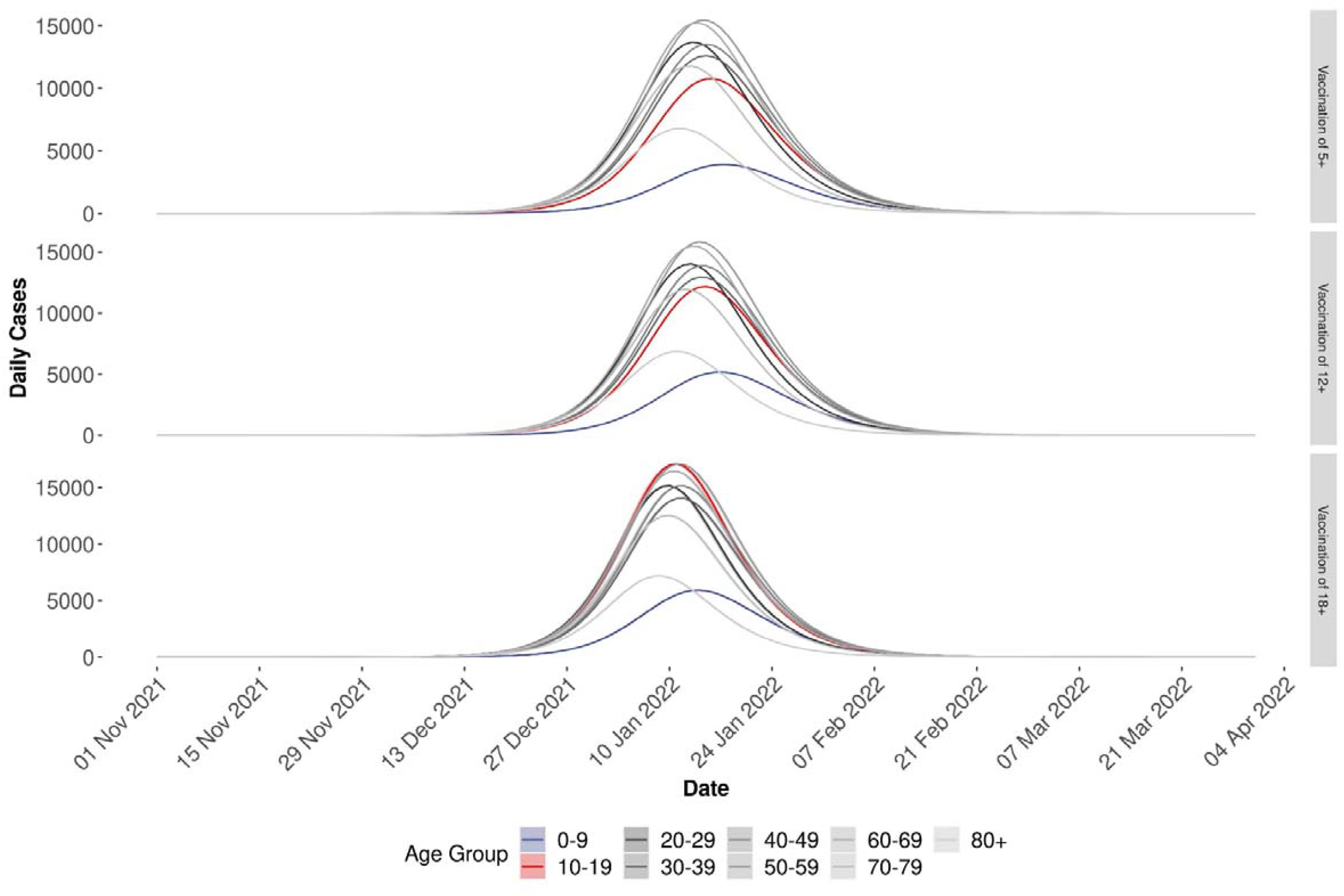
Daily cases by age group by vaccination scenario. Simulations were run from 22 June 2021 until 31 March 2022.

In our main analysis, in which we assumed no waning of vaccine effectiveness, we found overall reductions of 5.7% (4.4%, 6.9%) of cases, 2.0% (0.7%, 3.2%) of hospital admissions, and 1.7% (0.6%, 2.8%) of IC admissions when adolescents were included in the vaccination program (12+) compared to vaccinating only adults (18+) (Table 1). This corresponded to absolute reductions of 50806 (47357, 55675) cases, 21 (19, 23) hospital admissions, and 5 (5, 5) IC admissions compared to vaccination of adults only. Percent reductions were largest in the 10-19 age group with reductions of 22.5% (21.4%, 23.4%) of cases, 22.5% (21.4%, 23.5%) of hospital admissions, and 36.0% (35.3%, 36.7%) of IC admissions.

**Table 1.** Cumulative sum, absolute difference, and percent difference of modelled outcomes by vaccination scenario (18+, 12+, 5+) and age group. Vaccination in 18+ is used as the reference for absolute and percent difference. Absolute difference is calculated as outcome in 12+ or 5+ minus outcome in 18+. We assume vaccine efficacy does not wane. IC = intensive care, CI = confidence interval.

| Age Group | Outcome | Cumulative Sum (95% CI) |  |  | Absolute Difference (95% CI) |  | Percent (%) Difference (95% CI) |  |
| --- | --- | --- | --- | --- | --- | --- | --- | --- |
|  |  | Vaccination of 18+ | Vaccination of 12+ | Vaccination of 5+ | Vaccination of 12+ | Vaccination of 5+ | Vaccination of 12+ | Vaccination of 5+ |
| Overall | Cases | 2529076<br>(2455767, 2602490) | 2384939<br>(2347519, 2422402) | 2308430<br>(2272414, 2344496) | -144137<br>(-108249, -180089) | -220646<br>(-183353, -257995) | -5.7<br>(-6.9, -4.4) | -8.7<br>(-9.9, -7.5) |
|  | Hospital | 44806 | 43923 | 43351 | -883 | -1454 | -2.0 | -3.2 |
|  | Admissions | (43539, 46074) | (43244, 44602) | (42684, 44020) | (-294, -1472) | (-855, -2055) | (-3.2, -0.7) | (-4.5, -2.0) |
|  | IC | 9506 | 9345 | 9280 | -161 | -226 | -1.7 | -2.4 |
|  | Admissions | (9266, 9745) | (9215, 9474) | (9152, 9409) | (-51, -271) | (-114, -337) | (-2.8, -0.6) | (-3.5, -1.2) |
| 0-9 | Cases | 131363<br>(127826, 134910) | 120819<br>(119058, 122583) | 92343<br>(91000, 93691) | -10544<br>(-12326, -8768) | -39020<br>(-41219, -36826) | -8.0<br>(-9.1, -6.9) | -29.7<br>(-30.6, -28.8) |
|  | Hospital | 1191 | 1096 | 838 | -96 | -353 | -8.0 | -29.7 |
|  | Admissions | (1160, 1222) | (1080, 1111) | (826, 850) | (-111, -80) | (-373, -334) | (-9.1, -6.9) | (-30.5, -28.8) |
|  | IC | 0 | 0 | 0 | 0 | 0 | -- | -- |
|  | Admissions | (0, 0) | (0, 0) | (0, 0) | (0, 0) | (0, 0) | -- | -- |
| 10-19 | Cases | 350682<br>(340327, 361047) | 271719<br>(267489, 275956) | 245960<br>(242183, 249747) | -78963<br>(-85091, -72838) | -104721<br>(-111300, -98144) | -22.5<br>(-23.4, -21.4) | -29.9<br>(-30.8, -28.8) |
|  | Hospital | 144 | 112 | 101 | -32 | -43 | -22.5 | -39.8 |
|  | Admissions | (140, 148) | (110, 113) | (100, 103) | (-35, -30) | (-46, -40) | (-23.5, -21.4) | (-30.8, -28.9) |
|  | IC | 24 | 15 | 13 | -9 | -11 | -36.0 | -46.2 |
|  | Admissions | (23, 24) | (15, 15) | (12, 13) | (-9, -8) | (-11, -10) | (-36.7, -35.3) | (-46.8, -45.7) |
| > 19 | Cases | 2047032<br>(1987615, 2106533) | 1992402<br>(1960792, 2023862) | 1970127<br>(1939231, 2001057) | -54630<br>(-82671, -26642.7) | -76905<br>(-105476, -48384) | -2.7<br>(-3.9, -1.3) | -3.8<br>(-5.0, -2.4) |
|  | Hospital | 43470 | 42716 | 42413 | -755 | -1058 | -1.7 | -2.4 |
|  | Admissions | (42329, 44704) | (42051, 43378) | (41758, 43067) | (-1326, -184) | (-1636, -481) | (-3.0, -0.4) | (-3.7, -1.1) |
|  | IC | 9482 | 9330 | 9267 | -153 | -215 | -1.6 | -2.3 |
|  | Admissions | (9243, 9721) | (9200, 9459) | (9139, 9396) | (-263, -43) | (-326, 104) | (-2.7, -0.5) | (-3.3, -1.1) |

When vaccination was extended further to include 5-11 year olds, we saw greater reductions in disease outcomes compared to only extending vaccination to adolescents (12+). When children were included in the vaccination program, we observed an 8.7% (7.5%, 9.9%) reduction in cases, 3.2% (2.0%, 4.5%) reduction in hospital admissions, and 2.4% (1.2%, 3.5%) reduction in IC admissions in the entire population compared to vaccination in adults only (Table 1). These percent reductions corresponded to absolute reductions of 220646 (183353, 257995) cases, 1454 (855, 2055) hospital admissions, and 226 (114, 337) IC admissions. As with the 12+ scenario, benefits were larger within the age groups included in the vaccination program extension. Percent reductions in cases and hospital admissions in the 0-9 age group were 29.7% (28.8%, 30.6%) and 29.7% (28.8%, 30.5%), respectively in the 5+ scenario compared to the 18+ scenario. Within the 10-19 age group we observed reductions of 29.9% (28.8%, 30.8%) cases, 39.8% (28.9%, 30.8%) hospital admissions, and 46.2% (45.7%, 46.8%) IC admissions in the 5+ scenario compared to the 18+ scenario.

When looking all 0-19 year olds, under the 12+ scenario, we saw absolute reductions of 89507 (81606, 97417) cases, 128 (110, 146) hospital admissions, and 9 (8, 9) IC admissions compared to vaccination in adults only. We saw larger absolute reductions in disease outcomes in 0-19 year olds when everyone 5 years and above was vaccinated, specifically absolute reductions of 143741 (134970, 152519) cases, 396 (375, 419) hospital admissions, and 11 (10, 11) IC admissions compared to vaccination in adults only (Table 1).

### Sensitivity Analyses

To determine the robustness of our results, we performed sensitivity analyses in which we assumed vaccine-induced immunity waned completely after 1 year. Firstly, we observed a larger and earlier epidemic compared to when no waning was assumed (Figure 3) and that cases were driven by the oldest age groups, who were vaccinated first, regardless of vaccination strategy (Figure S4).

**Figure 3.**
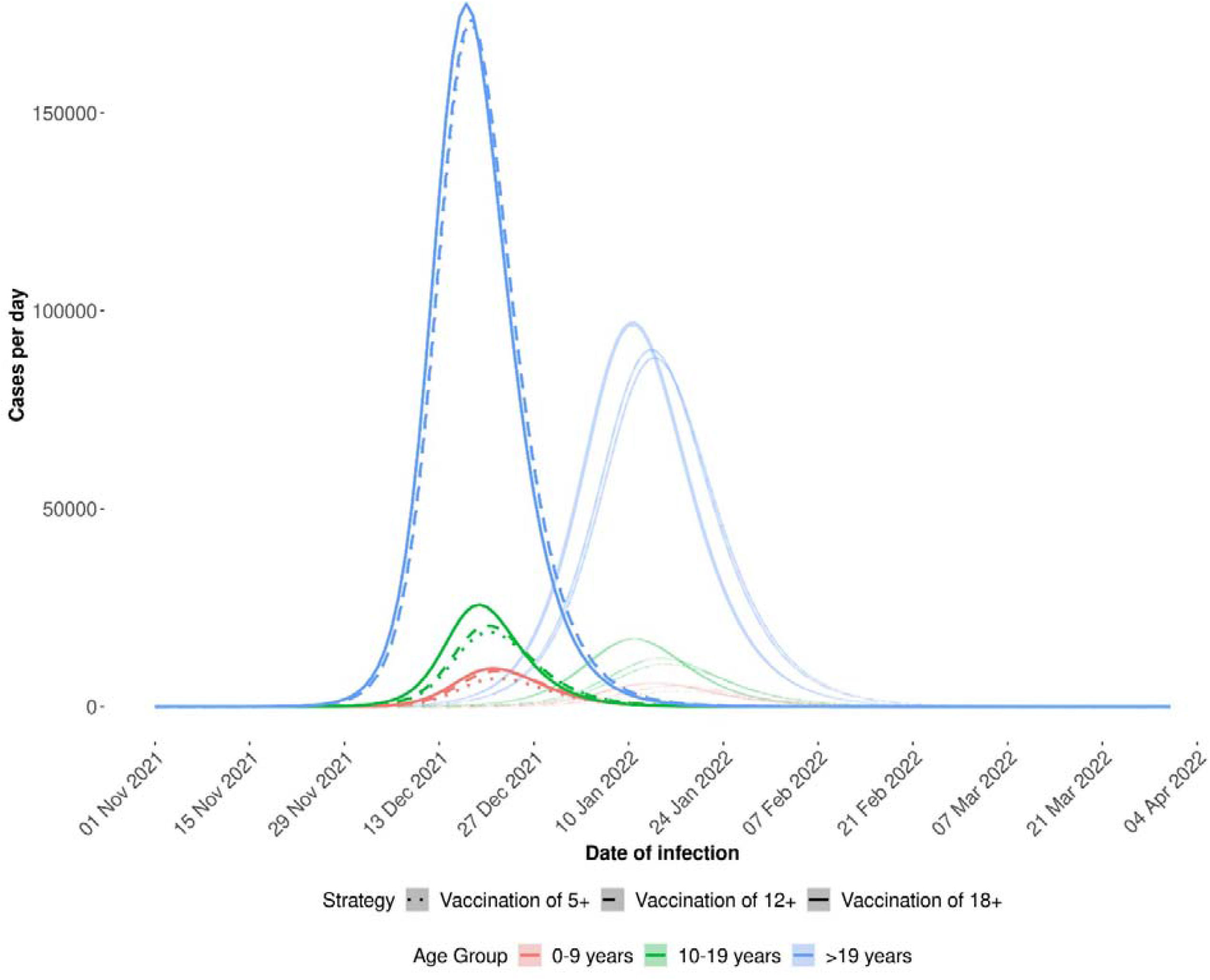
Daily cases by vaccination scenario and age group in the absence (faded lines) and presence of waning vaccine-induced immunity. Simulations were run from 22 June 2021 until 31 March 2022. Control measures are relaxed on 1 November 2021.

Secondly, we observed smaller percent reductions in disease outcomes by extending vaccination to adolescents and children compared to when vaccination did not wane. Overall we observed percent reductions of 2.7% (0.9%, 4.3%) cases, 0.5% (−1.3%, 2.2%) hospital admissions, and 0.6% (−0.9%, 1.9%) IC admissions under the 12+ scenario. We observed percent reductions of 4.7% (2.9%, 6.3%) cases, 1.2% (0.6%, 2.9%) hospital admissions, and 0.8% (0.6%, 2.1%) IC admissions under the 5+ scenario (Table 2). Despite smaller percent reductions when waning vaccine-induced immunity was assumed, extending vaccination beyond adults resulted in large reductions of disease outcomes. As in our main analysis, reductions in disease outcomes were highest in the age groups that were included in the vaccination program extension (Table 2).

**Table 2.** Cumulative sum, absolute difference, and percent difference of modelled outcomes by vaccination scenario (18+, 12+, 5+) and age group. with and without vaccination in 12 – 17 year olds. Vaccination in 18+ is used as the reference for absolute and percent difference. Absolute difference is calculated as outcome in 12+ or 5+ minus outcome in 18+18+ - outcome in 12+. We assume vaccine efficacy wanes completely after 1 year. IC = intensive care, CI = confidence interval.

| Age Group | Outcome | Cumulative Sum (95% CI) |  |  | Absolute Difference (95% CI) |  | Percent (%) Difference (95% CI) |  |
| --- | --- | --- | --- | --- | --- | --- | --- | --- |
|  |  | Vaccination of 18+ | Vaccination of 12+ | Vaccination of 5+ | Vaccination of 12+ | Vaccination of 5+ | Vaccination of 12+ | Vaccination of 5+ |
| Overall | Cases | 3173616<br>(2994529, 3352088) | 3087975<br>(2968635, 3206806) | 3024027<br>(2907191, 3140369) | -85641<br>(-145281, -25895) | -149589<br>(-211718, -87338) | -2.7<br>(-4.3, -0.9) | -4.7<br>(-6.3, -2.9) |
|  | Hospital | 60027 | 59708 | 59280 | -318 | -747 | -0.5 | -1.2 |
|  | Admissions | (56639, 63406) | (57380, 62031) | (56965, 61588) | (-1375, 740) | (-1818, 326) | (-2.2, 1.3) | (-2.9, 0.6) |
|  | IC | 10117 | 10061 | 10035 | -56 | -82 | -0.6 | -0.8 |
|  | Admissions | (9660, 10574) | (9746, 10375) | (9722, 10349) | (-199, 86) | (-225, 61) | (-1.9, 0.9) | (-2.1, 0.6) |
| 0-9 | Cases | 160841<br>(153090, 168544) | 153896<br>(148840, 158918) | 121396<br>(117402, 125374) | -6945<br>(-9626, -4250) | -39445<br>(-43170, -35688) | -4.3<br>(-5.7, -2.8) | -24.5<br>(-25.6, -23.3) |
|  | Hospital | 1458 | 1395 | 1101 | -63 | -357 | -4.3 | -24.5 |
|  | Admissions | (1391, 1525) | (1352, 1439) | (1067, 1136) | (-86, -40) | (-389, -325) | (-5.6, -2.9) | (-23.3, 25.5) |
|  | IC | 0 | 0 | 0 | 0 | 0 | -- | -- |
|  | Admissions | (0, 0) | (0, 0) | (0, 0) | (0, 0) | (0, 0) |  |  |
| 10-19 | Cases | 378810<br>(358010, 399564) | 322359<br>(310778, 333877) | 302596<br>(291884, 313243) | -56451<br>(-65686, -47232) | -76214<br>(-86321, -66126) | -14.9<br>(-16.4, -13.2) | -20.1<br>(-21.6, -18.5) |
|  | Hospital | 156 | 132 | 124 | -23 | -31 | -14.9 | -20.1 |
|  | Admissions | (148, 164) | (128, 137) | (120, 129) | (-27, -20) | (-35, -27) | (-16.3, -13.3) | (-21.5, -18.6) |
|  | IC | 25 | 17 | 14 | -8 | -11 | -33.3 | -42.9 |
|  | Admissions | (24, 26) | (16, 17) | (14, 15) | (-9, -8) | (-11, -10) | (-33.7, -32.2) | (-43.3, -41.7) |
| > 19 | Cases | 2633964.<br>(2483429, 2783980) | 2611719<br>(2509016, 2714011) | 2600035<br>(2497905, 2701753) | -22245<br>(-69969, 25587) | -33930<br>(-82228, 14476) | -0.8<br>(-2.5, 1.0) | -1.3<br>(-3.0, 0.6) |
|  | Hospital | 58413 | 58180 | 58054 | -232 | -359 | -0.4 | -0.6 |
|  | Admissions | (55100, 61717) | (55900, 60455) | (55779, 60324) | (-1262, 800) | (-1393, 678) | (-2, 1.5) | (-2.3, 1.2) |
|  | IC | 10092 | 10044 | 10021 | -48 | -71 | -0.5 | -0.7 |
|  | Admissions | (9636, 10548) | (9730, 10358) | (9708, 10334) | (-190, 94) | (-214, 72) | (-1.8, 1.0) | (-2.0, 0.7) |

## Discussion

We quantified the impacts of extending vaccination to adolescents and children against COVID-19 on disease outcomes. We first quantify the impacts of vaccinating 12-17 year olds against COVID-19. However, this policy decision (extending vaccination to healthy adolescents) has already been made in many countries. Future policy will focus on further extension to vaccination programs, such as vaccination of 5-11 year olds [62]. Therefore, we also quantify the impacts of extending COVID-19 vaccination programs further to children aged 5-11 years.

We found that upon the release of all non-pharmaceutical control measures, an epidemic occurred regardless of vaccination strategy. We also found that due to the increased transmissibility of the Delta variant and reduced vaccine effectiveness (particularly against infection), the reductions in disease outcomes in our updated analysis were smaller than our original analysis which our assumed transmission rate and vaccine effectiveness were consistent with the Alpha variant (Table S5). We found that vaccinating 75% of 12-17 year olds resulted in reductions of 5.7% (4.4%, 6.9%) of cases, 2.0% (0.7%, 3.2%) of hospital admissions, and 1.7% (0.6%, 2.8%) of IC admissions in the entire population, but that percent reductions in disease outcomes were larger in the 10-19 age group age group (22.5% of cases, 22.5% of hospital admissions, and 36.0% of IC admissions). When vaccination was extended further to include 5-11 year olds, we observed overall reductions of 8.7% (7.5%, 9.9%) of cases, 3.2% (2.0%, 4.5%) of hospital admissions, and 2.4% (1.2%, 3.5%) of IC admissions. Percent reductions were higher in the 0-9 and 10-19 age groups. Our results were robust to assumptions about waning vaccine-induced immunity, but the benefits of vaccinating adolescents and children were smaller if vaccine protection was assumed to wane.

Our results highlight that vaccinating 5-11 and/or 12-17 year olds is not sufficient to avert a future epidemic without the re-introduction of non-pharmaceutical interventions. However, in the simulations presented here, we assume that after control measures are relaxed they are never re-implemented. In reality, if cases and severe disease outcomes rise again, control measures would be re-implemented. Therefore, the averted number of disease outcomes reported here should be used for comparison of the vaccination strategies only and not as a prediction of the actual number of outcomes that will occur. In a more realistic scenario where measures are re-implemented to control rising infections we would expect the absolute number of averted disease outcomes to be smaller, but the indirect benefits of vaccinating children and adolescents to be higher.

When we looked at disease outcomes by age group we saw that adolescents had one of the highest incidences of cases when not included in the vaccination program (Figure 2, 18+ panel, red line); however, incidence among adolescents fell below other age groups when adolescents were vaccinated (Figure 2, 5+ and 12+ panels, red line). In future, it may be necessary to implement control measures to avoid a large rise in SARS-CoV-2 infections and severe disease outcomes. Physical distancing measures will be most effective if they are targeted at age groups that contribute most to further spread, i.e, the age groups with the highest incidence of infection [37]. That implies that closure of secondary schools, which reduces contacts among the age group with the highest incidence of infection in the absence of an adolescent vaccination program, will be a very effective non-pharmaceutical control measure. However, the negative effects of school closures have been well documented [35-37, 63]. Thus, an additional benefits of vaccinating adolescents, beyond reducing infections, is limiting quarantine for students and school closures, and minimizing education disruptions [16]. If 12–17 year olds are vaccinated, they are less likely to be the age group with the highest incidence, and there is no obvious need for closures of secondary schools. Public health policy that aims to minimize school closures should be a priority [64].

The work presented here has some limitations. First, we do not account for population heterogeneity beyond 10-year age groups; therefore, we do not account for additional sources of heterogeneity such as individuals with differing levels of risk (due to factors other than age, e.g., co-morbidities) and geographic clustering in vaccination coverage or naturally-acquired immunity. Therefore, these results represent the average impacts of vaccinating adolescents in the population, but precise benefits may vary in subgroups of the population. Second, we assume that once individuals recover from infection, they cannot be re-infected. There is evidence that re-infection with SARS-CoV-2 [65] may be possible; therefore, we may be underestimating the number of infections. We also assume a final vaccination coverage of 75% in children and adolescents, which is consistent with the percent of adolescents willing to be vaccinated in The Netherlands [66]; however, this may be optimistic. If true vaccination coverage in children and adolescents is lower, then the impacts of vaccinating these groups will also be lower. Finally, we assume there is no change in the probability of severe disease upon infection over time or between variants. There is evidence that suggests increased disease severity after infection with the Delta variant [67]; therefore, we may be underestimating the number of severe infections.

In conclusion, our results highlight the benefits to extending COVID-19 vaccination programs to children and adolescents to reduce infections and severe outcomes, both in children and adolescents themselves and in the population as a whole. However, extending vaccinating alone will not prevent a future epidemic. Therefore, additional control measures may need to be implemented to prevent a large increase in hospital and IC admissions. While the results presented here are based on population characteristics and the COVID-19 vaccination program in The Netherlands, they may provide valuable insights for other countries who have not yet extended vaccination beyond adults. Furthermore, a reduction of infections in children and/or adolescents in concert with additional control measures may have the added benefit of reducing the need for school closures in the event of an epidemic. After the education disruptions primary and secondary aged children have endured throughout the pandemic, policies that will limit further disruption should be considered with priority.

## Supplemental Material

### Methods

We developed a deterministic age-structured compartmental susceptible-exposed-infectious-recovered (SEIR) model extended to include states for severe disease outcomes and vaccination status. The population is partitioned into 10-year age groups (0-9, 10-19, …, 70-79, 80+). Within each age group we further partition the population into those who are unvaccinated, vaccinated with 1 dose, or vaccinated with 2 doses and then finally into disease states: susceptible (S), infected but not yet infectious (E), infectious (I), hospitalized (H), in intensive care (IC), return to the hospital ward after intensive care (HIC), recovered (R), and dead (D) (Figure S1).

**Figure S1.**
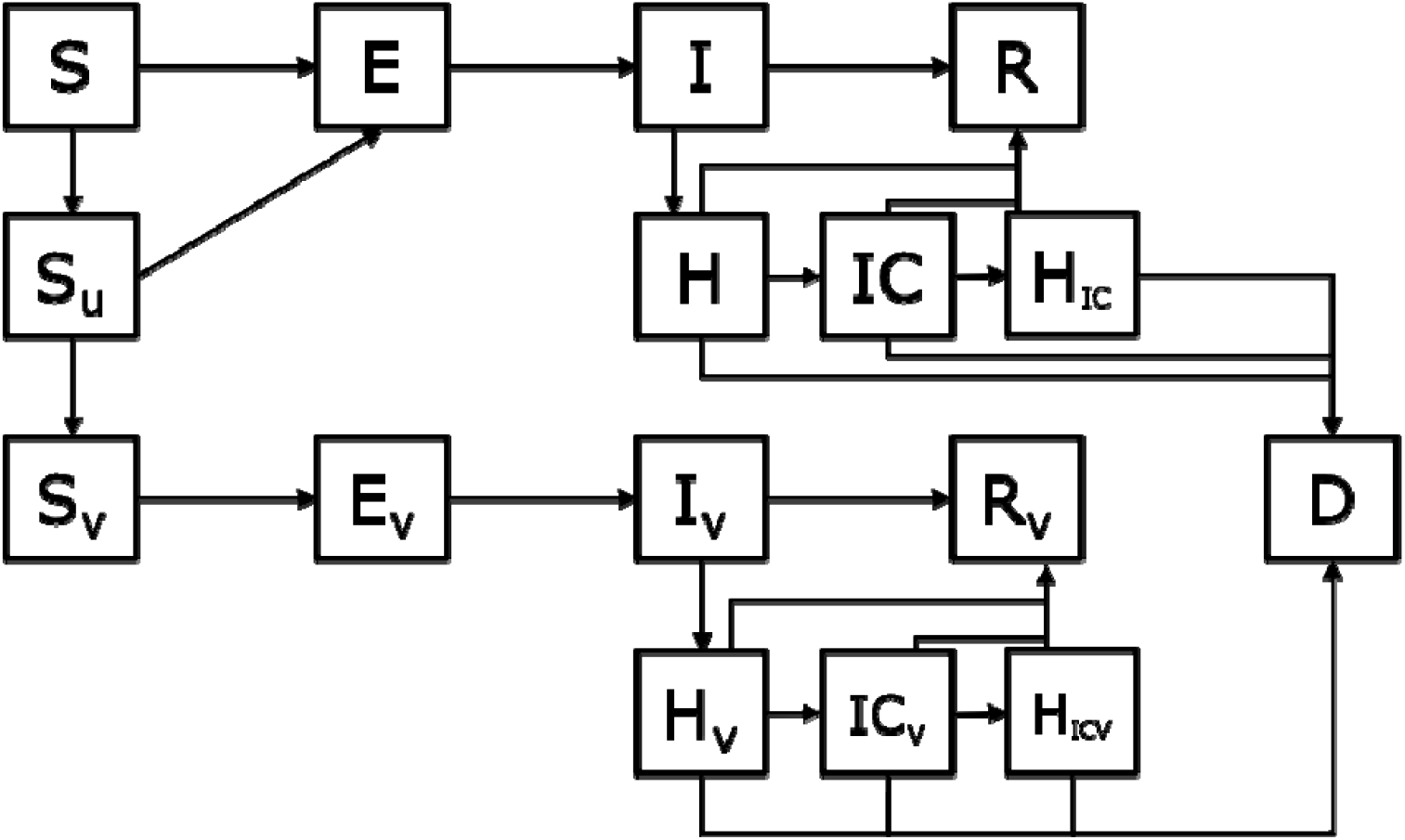
Basic conceptual model diagram. This diagram does not include the additional states after the second dose of vaccination or the age structure in the model. S = susceptible, E = exposed, I = Infectious, R = Recovered, H = hospitalized, IC = In intensive care, HIC = return to the hospital ward following treatment in IC, Su = vaccinated, but not yet protected, D = dead. States with subscript V indicate individuals who are vaccinated and protected by vaccination. This model assumes the “leaky” vaccine protection, so vaccinated and protected individuals can still be infected, hospitalized, etc. but at a reduced rate.

When a person is vaccinated, they first enter a hold state where they are vaccinated, but not yet (fully) protected (Shold1d or Shold2d). After a delay period, they enter the vaccinated and protected state for the dose they have received (Sv1d or Sv2d). We assume only susceptible individuals are vaccinated. We use the model to determine the number of daily cases, hospital admissions, and IC admissions under different vaccination scenarios. The mathematical equations for determining each outcome and the differential model equations are shown below. In the model equations, bold capital letters refer to matrices, bold lower case letters indicate vectors, and plain text lower case letters indicate scalars. Since model compartments are denoted with capital letters, but are vectors due to the age-stratification of the model, we denote them with a half arrow above the compartment symbol, e.g.,. Parameter definitions and values are shown in Table S3 and Table S4.

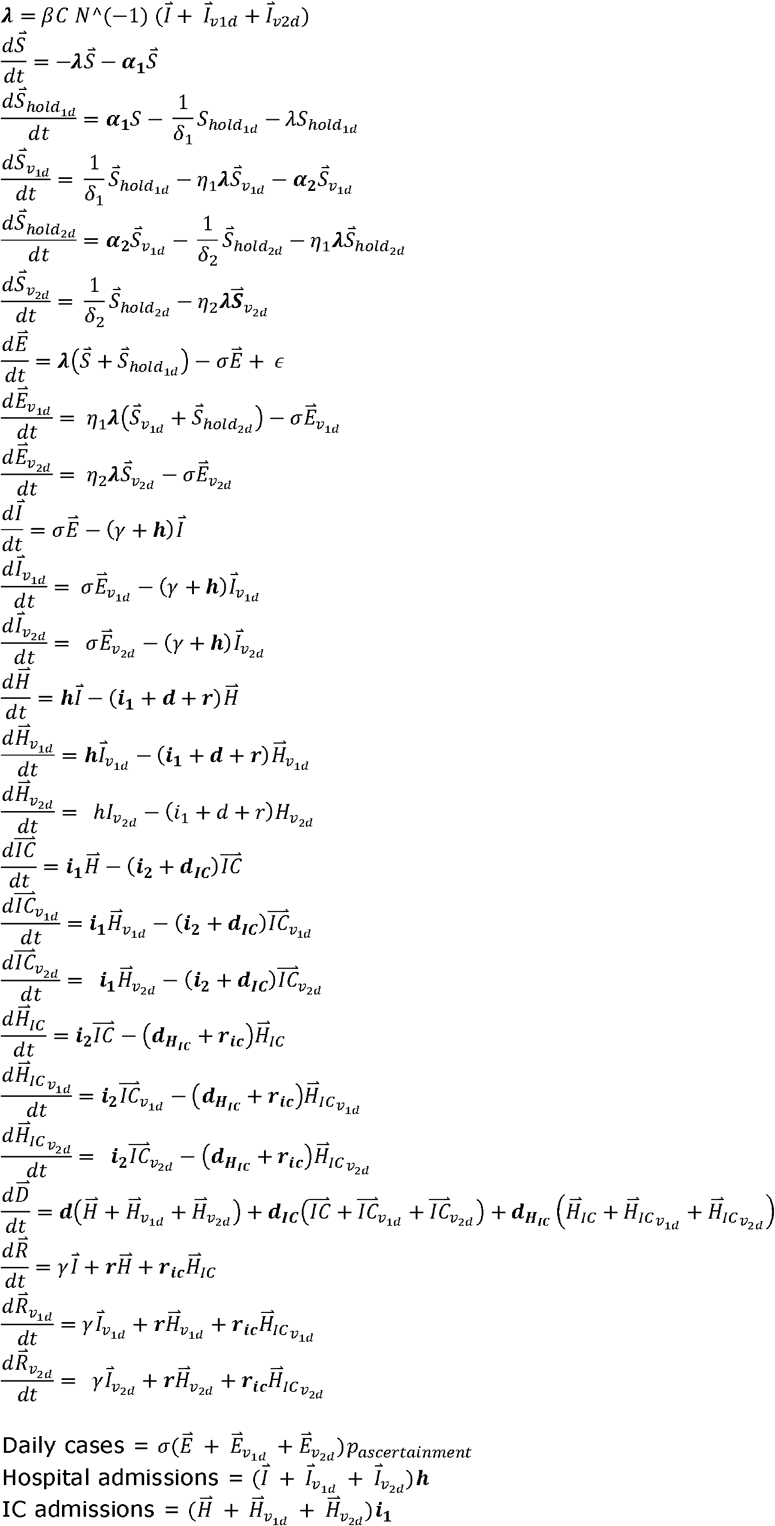

The model is designed to incorporate a single vaccine product with a 2-dose regimen that 1) reduces susceptibility to infection, 2) reduces risk of hospitalization if a vaccinated individual is infected, and 3) reduces risk of infecting others (transmission) if a vaccinated person is infected. However, since there are more than one vaccine products currently licensed for use against COVID-19 in The Netherlands (the vaccines made by Pfizer/BioNTech, Moderna, AstraZeneca, and Janssen), we incorporate different vaccine products by taking the daily weighted average of the number of people with each vaccine product (and dose), the corresponding delay to protection of each vaccine product, and the vaccine effectiveness against each outcome (Table S1). Janssen is incorporated by using zero for the number of second doses at all time points. The weighted average of the VE can be expressed as:

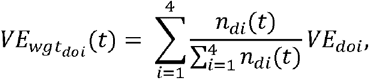

where *n*_*di*_(*t*) is the number of vaccines given as dose *d* (d = 1, 2) from vaccine product *i* (i = 1, 2, 3, 4) at time t and *VE*_*doi*_ is the VE against outcome *o* (*o* = infection, hospitalization, transmission), for dose *d*, from vaccine product *i*.

When VE is assumed to wane, we include the amount of waning as a logistic function parameterized so that after 6 months vaccine-induced protection is reduced by 50% and reduced by 100% after 1 year. The amount of waning is calculated at a given time since vaccination as

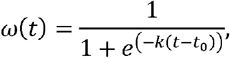

where k is the logistic growth rate (here, k = 0.03), t is the time since vaccination in days, and t_0_ is the time point (in days) where 50% reduction occurs (here, t_0_=180). Then at each timepoint, VE with waning (VE_ω_ (t))is calculated as

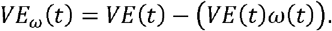

If(*VE*(*t*)*ω* (*t*) *VE*_*ω*_ (*t*)= 0

Rate of vaccination for each day, vaccine product, and dose for each age group is a model input. Increase of vaccination coverage over time was based on weekly data of allocated vaccines (up to 16 June 2021) or projected available vaccines (after 16 June 2021) by vaccine type, dose number, and target group (split up between healthcare workers, residents in institutions, chronically ill, and, if not part of one of the aforementioned groups, by age in 10-years age-bands) [68]. Projected final vaccination coverages were assumed at 90% for ≥70-year-olds and residents in institutions, 85% for 60-to 69-years-olds, health care workers and high-risk individuals, and 75% for others below 60 years of age. It was assumed that available vaccines were administered immediately until a target group reached the final coverage.

To account for the seasonal pattern of SARS-CoV-2 whereby, transmission is lower in summer and higher in winter, we define the transmission rate at time *t, β*(*t*), as a sinusoidal function of seasonality [6]:

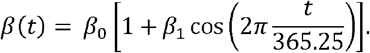

*β*_0_ is a baseline (non-seasonal) transmission rate, β_l_ is the amplitude of seasonal forcing, and *t* is the day of the year. We assume β_l_ = 0.14. The baseline transmission rate β_0_ and initial conditions for forward simulations are estimated by fitting the model to daily cases from the national notification database Osiris from 01 January 2020 to 22 June 2021, when vaccination in 12-17 year olds began in The Netherlands (Figure S2). The model is fitted to data piecewise to correspond with the correct contact patterns associated with different non-pharmaceutical interventions within each time window (Table S2). We directly estimate effective reproduction number (R_t_) within each time window using maximum likelihood estimation. We assume daily cases follow a negative binomial distribution with mean μ and overdispersion parameter *φ*. Estimates of R_t_ are converted to transmission rate by: 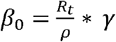, where γ is the inverse of the infectious period and ρ is the dominant eigen value of the product of the diagonal matrix of the number of susceptibles and the transmission matrix. The transmission matrix is determined by multiplying the rows and columns of the contact matrix by estimates of the relative susceptibility and infectiousness of each age group compared to the 0-9 years age group (Table S4).

The amplitude e of seasonal forcing β_l_ is estimated by linear regression:

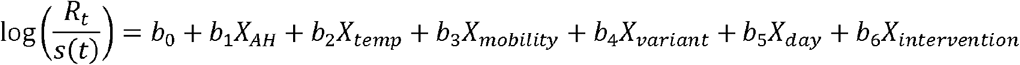

The response variable is the logarithm of the effective reproduction number *R*_*t*_ [69] divided by the fraction susceptible s(t). The fraction susceptible is estimated from the cumulative age-specific hospitalizations at time t divided by age-specific hospitalization rates, which were in turn estimated from seroprevalences after the first wave in The Netherlands [69]. The explanatory variables for seasonal changes are absolute humidity (X_AH_)and temperature in degrees Celsius (X_temp_) [70]. Additional explanatory variables that may affect *R*_*t*_ were also included: percent change in mobility from “workplace” and “retail and recreation” (X_mobility_)[71], increased transmissibility due to change in circulating virus variants (notably alpha and delta) (X_variant_)[72], day of the week (X_day_), and intervention period (X_intervention_). Intervention period is a variable indicating 2-week to 2-month time periods during the epidemic without change in control measures. New periods started 2020-03-13, 2020-05-06, 2020-06-01, 2020-07-01, 2020-09-01, 2020-09-29, 2020-10-26, 2020-12-21, 2021-01-23, 2021-02-08, 2021-03-01, 2021-04-26, 2021-05-17, 2021-06-05. The seasonality curve was extracted from the intercept and seasonal variable coefficients only, from which the amplitude was determined by fitting a sinusoidal curve.

**Figure S2.**
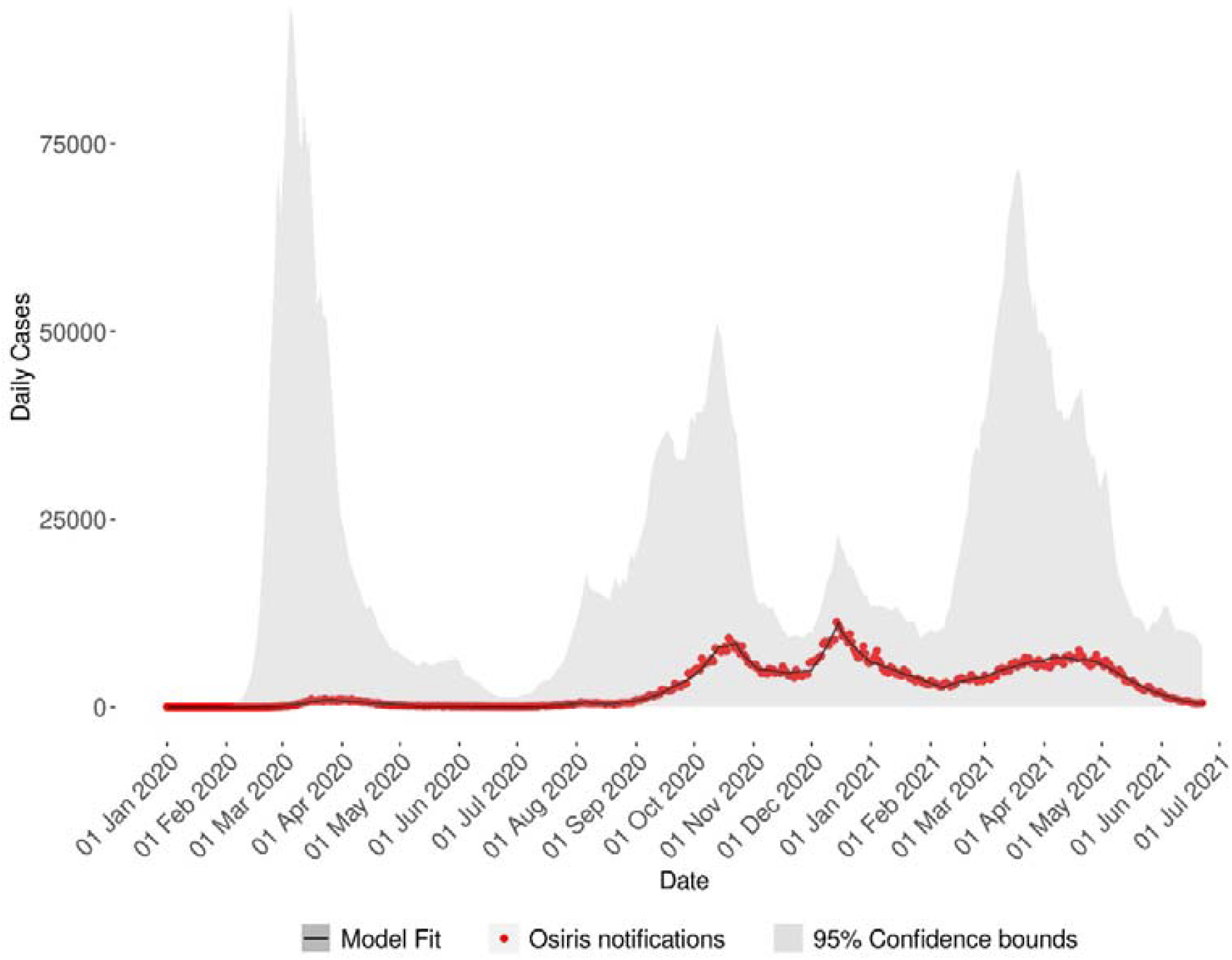
Fit to case notification data with 95% confidence bounds

**Figure S3.**
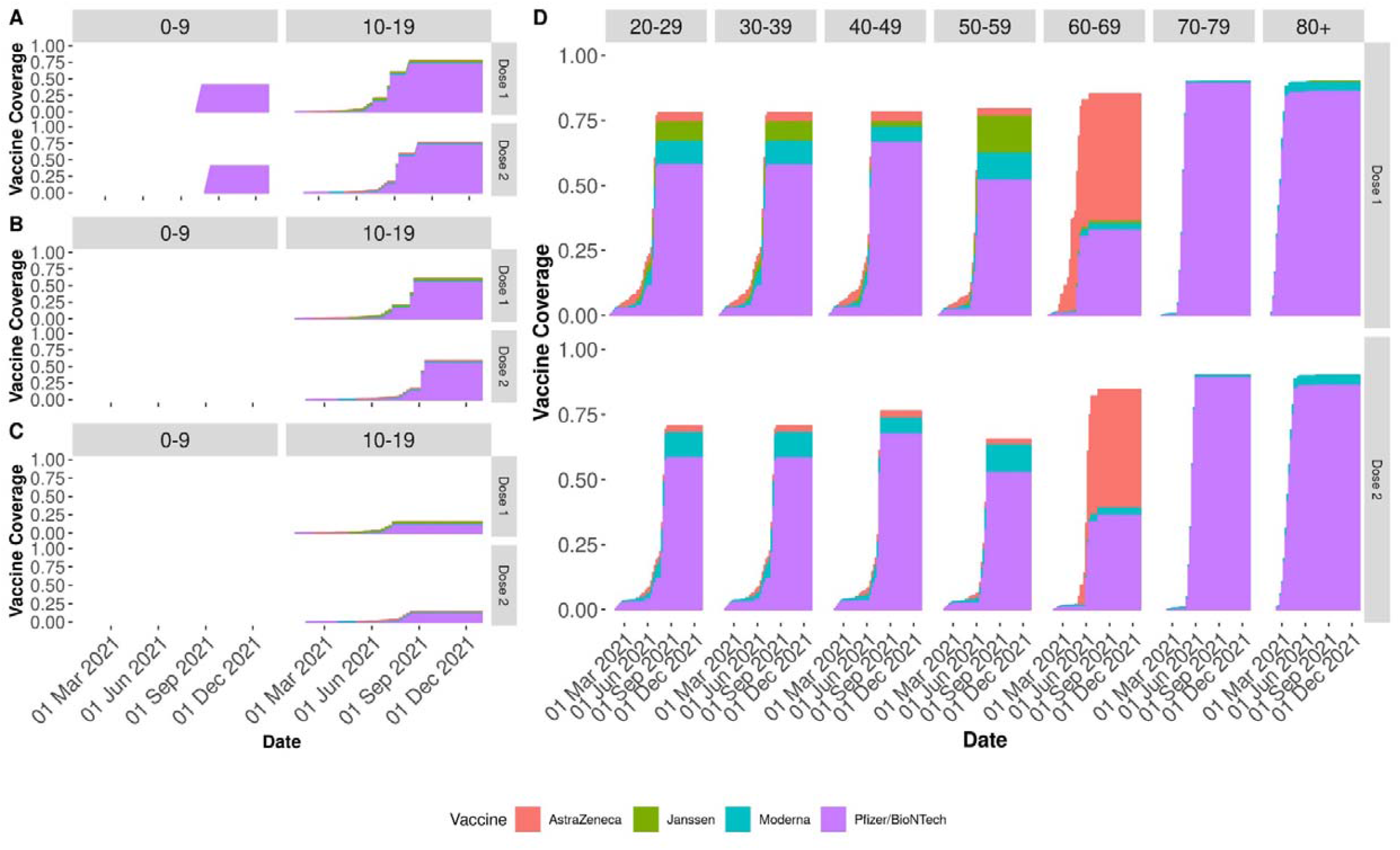
Vaccination coverage over time by dose, vaccine type, and age group for the different vaccination scenarios. A) vaccination coverage in 0-9 and 10-19 year old when everyone 5 years and above (5+) is vaccinated. B) vaccination coverage in 0-9 and 10-19 year old when everyone 12 years and above (12+) is vaccinated. C) vaccination coverage in 0-9 and 10-19 year old when everyone 18 years and above (18+) is vaccinated. D) vaccination coverage for adult age groups, (vaccination coverage in adults 20 years and above do not vary by vaccination scenario).

Our approach was used to inform Dutch COVID-19 policy regarding the vaccination of 12-17 year olds prior to the emergence of the Delta variant. For these simulations, forward simulations were performed from 22 June 2021 to 31 March 2022 with the initial conditions (i.e., the number of people in each compartment when the simulation begins) based on the last day of the model fitting. We assume the simulations begin with the same β_0_ that was estimated from the last fitted time window (05 June to 22 June) and with contact patterns estimated during June 2021. It was not yet known when the government would lift COVID-19 non-pharmaceutical interventions, therefore, when cases fell below 0.5 cases per 100,000 people, we assumed all non-pharmaceutical control measures were relaxed for the remainder of the simulation period. Therefore, we assume the contact patterns change to those estimated prior to the COVID-19 pandemic in April 2017. Further, once control measures are relaxed, we assume a transmission rate of β_0_=0.00059, corresponding to the basic reproduction number of the Alpha variant (R0=3.45).

**Table S1.**
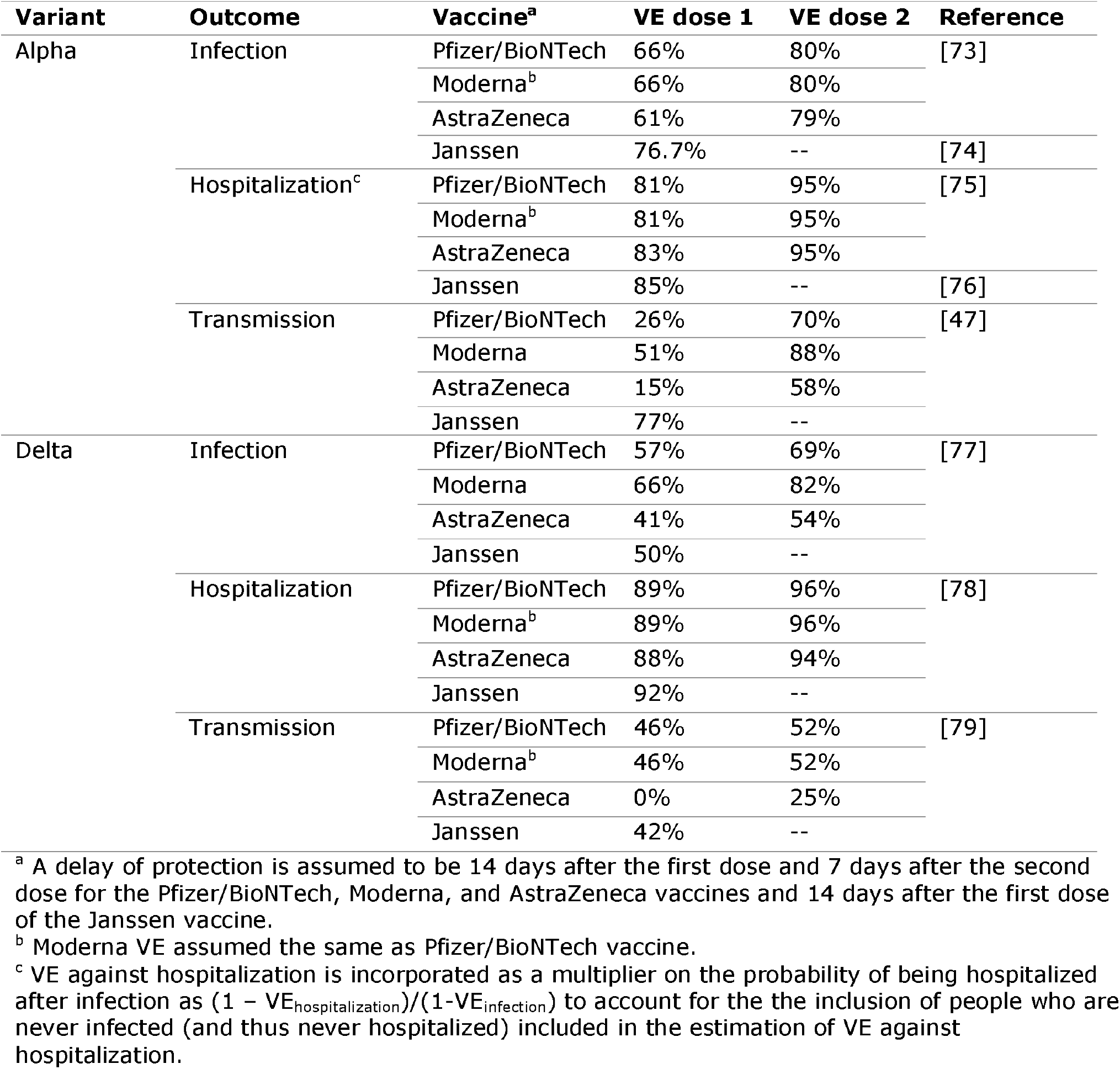
Vaccine effectiveness (VE) for each vaccine by dose based on observational studies.

**Table S2.**
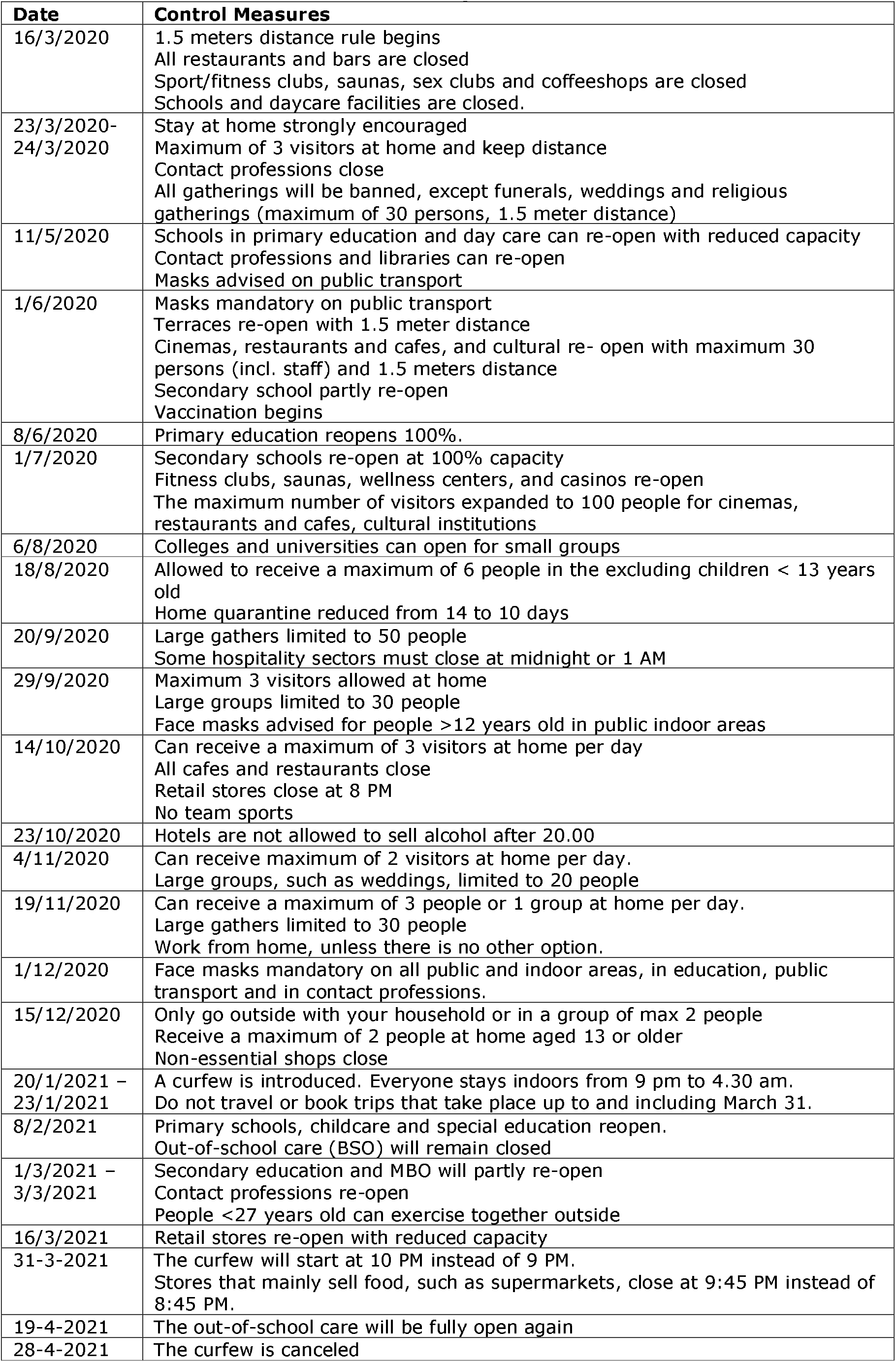
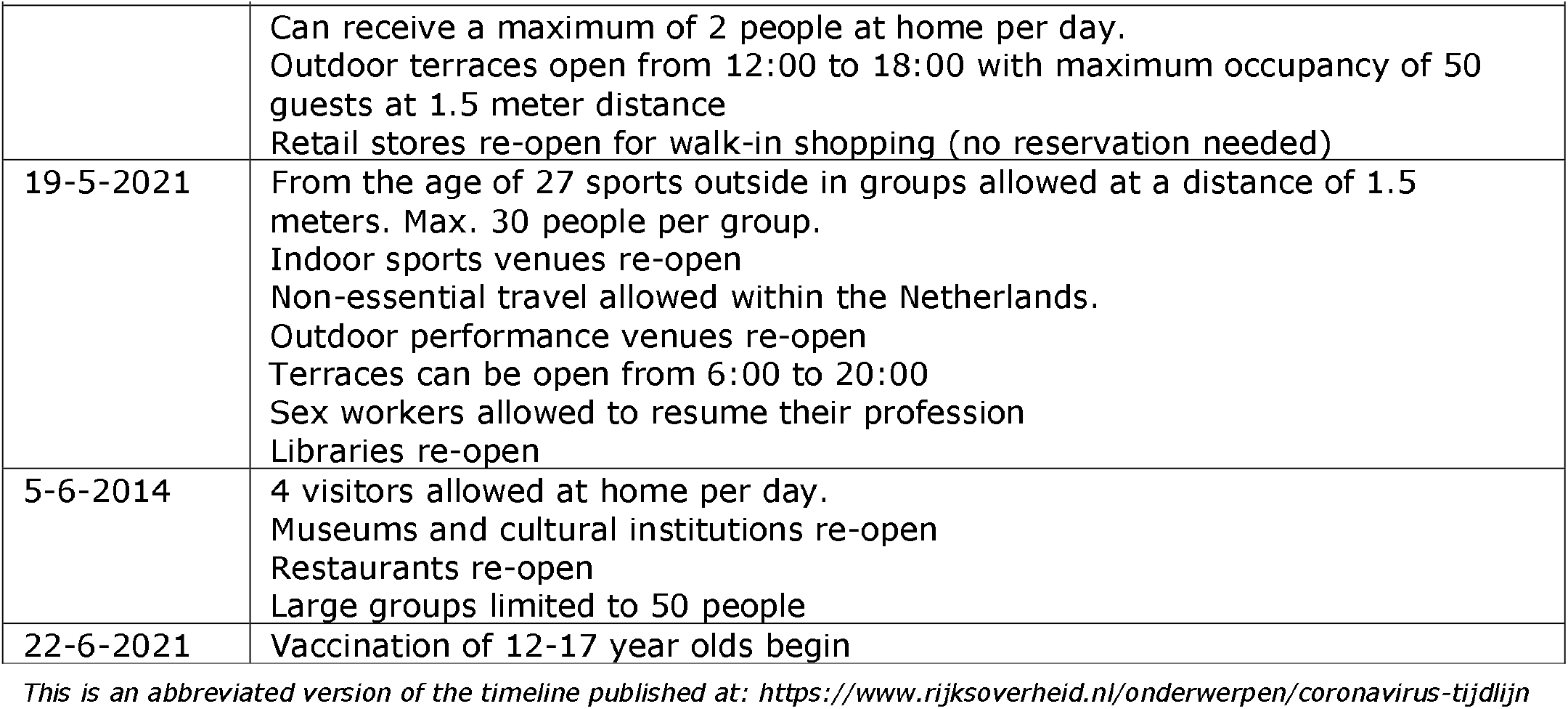
Timeline of measures and advice during the COVID-19 outbreak in the Netherlands.

| Date | Control Measures |
| --- | --- |
| 16/3/2020 | 1.5 meters distance rule begins<br>All restaurants and bars are closed<br>Sport/fitness clubs, saunas, sex clubs and coffeeshops are closed<br>Schools and daycare facilities are closed. |
| 23/3/2020-24/3/2020 | Stay at home strongly encouraged<br>Maximum of 3 visitors at home and keep distance<br>Contact professions close<br>All gatherings will be banned, except funerals, weddings and religious gatherings (maximum of 30 persons, 1.5 meter distance) |
| 11/5/2020 | Schools in primary education and day care can re-open with reduced capacity<br>Contact professions and libraries can re-open<br>Masks advised on public transport |
| 1/6/2020 | Masks mandatory on public transport<br>Terraces re-open with 1.5 meter distance<br>Cinemas, restaurants and cafes, and cultural re- open with maximum 30 persons (incl. staff) and 1.5 meters distance<br>Secondary school partly re-open<br>Vaccination begins |
| 8/6/2020 | Primary education reopens 100%. |
| 1/7/2020 | Secondary schools re-open at 100% capacity<br>Fitness clubs, saunas, wellness centers, and casinos re-open<br>The maximum number of visitors expanded to 100 people for cinemas, restaurants and cafes, cultural institutions |
| 6/8/2020 | Colleges and universities can open for small groups |
| 18/8/2020 | Allowed to receive a maximum of 6 people in the excluding children < 13 years old<br>Home quarantine reduced from 14 to 10 days |
| 20/9/2020 | Large gathers limited to 50 people<br>Some hospitality sectors must close at midnight or 1 AM |
| 29/9/2020 | Maximum 3 visitors allowed at home<br>Large groups limited to 30 people<br>Face masks advised for people >12 years old in public indoor areas |
| 14/10/2020 | Can receive a maximum of 3 visitors at home per day<br>All cafes and restaurants close<br>Retail stores close at 8 PM<br>No team sports |
| 23/10/2020 | Hotels are not allowed to sell alcohol after 20.00 |
| 4/11/2020 | Can receive maximum of 2 visitors at home per day.<br>Large groups, such as weddings, limited to 20 people |
| 19/11/2020 | Can receive a maximum of 3 people or 1 group at home per day.<br>Large gathers limited to 30 people<br>Work from home, unless there is no other option. |
| 1/12/2020 | Face masks mandatory on all public and indoor areas, in education, public transport and in contact professions. |
| 15/12/2020 | Only go outside with your household or in a group of max 2 people<br>Receive a maximum of 2 people at home aged 13 or older<br>Non-essential shops close |
| 20/1/2021 –<br>23/1/2021 | A curfew is introduced. Everyone stays indoors from 9 pm to 4.30 am.<br>Do not travel or book trips that take place up to and including March 31. |
| 8/2/2021 | Primary schools, childcare and special education reopen.<br>Out-of-school care (BSO) will remain closed |
| 1/3/2021 –<br>3/3/2021 | Secondary education and MBO will partly re-open<br>Contact professions re-open<br>People <27 years old can exercise together outside |
| 16/3/2021 | Retail stores re-open with reduced capacity |
| 31-3-2021 | The curfew will start at 10 PM instead of 9 PM.<br>Stores that mainly sell food, such as supermarkets, close at 9:45 PM instead of 8:45 PM. |
| 19-4-2021 | The out-of-school care will be fully open again |
| 28-4-2021 | The curfew is canceled |
|  | Can receive a maximum of 2 people at home per day.<br>Outdoor terraces open from 12:00 to 18:00 with maximum occupancy of 50 guests at 1.5 meter distance<br>Retail stores re-open for walk-in shopping (no reservation needed) |
| 19-5-2021 | From the age of 27 sports outside in groups allowed at a distance of 1.5 meters. Max. 30 people per group.<br>Indoor sports venues re-open<br>Non-essential travel allowed within the Netherlands.<br>Outdoor performance venues re-open<br>Terraces can be open from 6:00 to 20:00<br>Sex workers allowed to resume their profession<br>Libraries re-open |
| 5-6-2014 | 4 visitors allowed at home per day.<br>Museums and cultural institutions re-open<br>Restaurants re-open<br>Large groups limited to 50 people |
| 22-6-2021 | Vaccination of 12-17 year olds begin |

*This is an abbreviated version of the timeline published at: https://www.rijksoverheid.nl/onderwerpen/coronavirus-tijdlijn*

**Table S3.** Model parameters that do not vary with age.

| Parameter | Description | Value | Details |
| --- | --- | --- | --- |
| $\beta$ | Transmission rate | Varies over time | |
| $\sigma$ | Inverse of the latent period | 0.5/day | This results in a latent period of 2 days |
| $\gamma$ | Inverse of the infectious period | 0.5/day | This results in an infectious period of 2 days |
| $\lambda$ | Force of infection | Varies over time | |
| $\epsilon$ | Rate of case importation | 0.01/day | |
| $\alpha$ | Rate of vaccination with the first dose | This depends on the vaccine allocation schedule and varies over time | Calculated as a composite rate of multiple vaccines |
| $\alpha_2$ | Rate of vaccination with the second dose | This depends on the vaccine allocation schedule and varies over time | Calculated as a composite rate of multiple vaccines |
| $\delta_1$ | Delay to protection of first dose | See Table S1 | With multiple vaccines, the weighted average is used |
| $\delta_2$ | Delay to protection of second dose | See Table S1 | With multiple vaccines, the weighted average is used |
| $\eta$ | 1 – vaccine efficacy of first dose | See Table S1 | With multiple vaccines, the weighted average is used |
| $\eta_2$ | 1 – vaccine efficacy of second dose | See Table S1 | With multiple vaccines, the weighted average is used |
| $p_{\text{ascertainment}}$ | Case ascertainment rate | 0.3/day | |

**Table S4.**
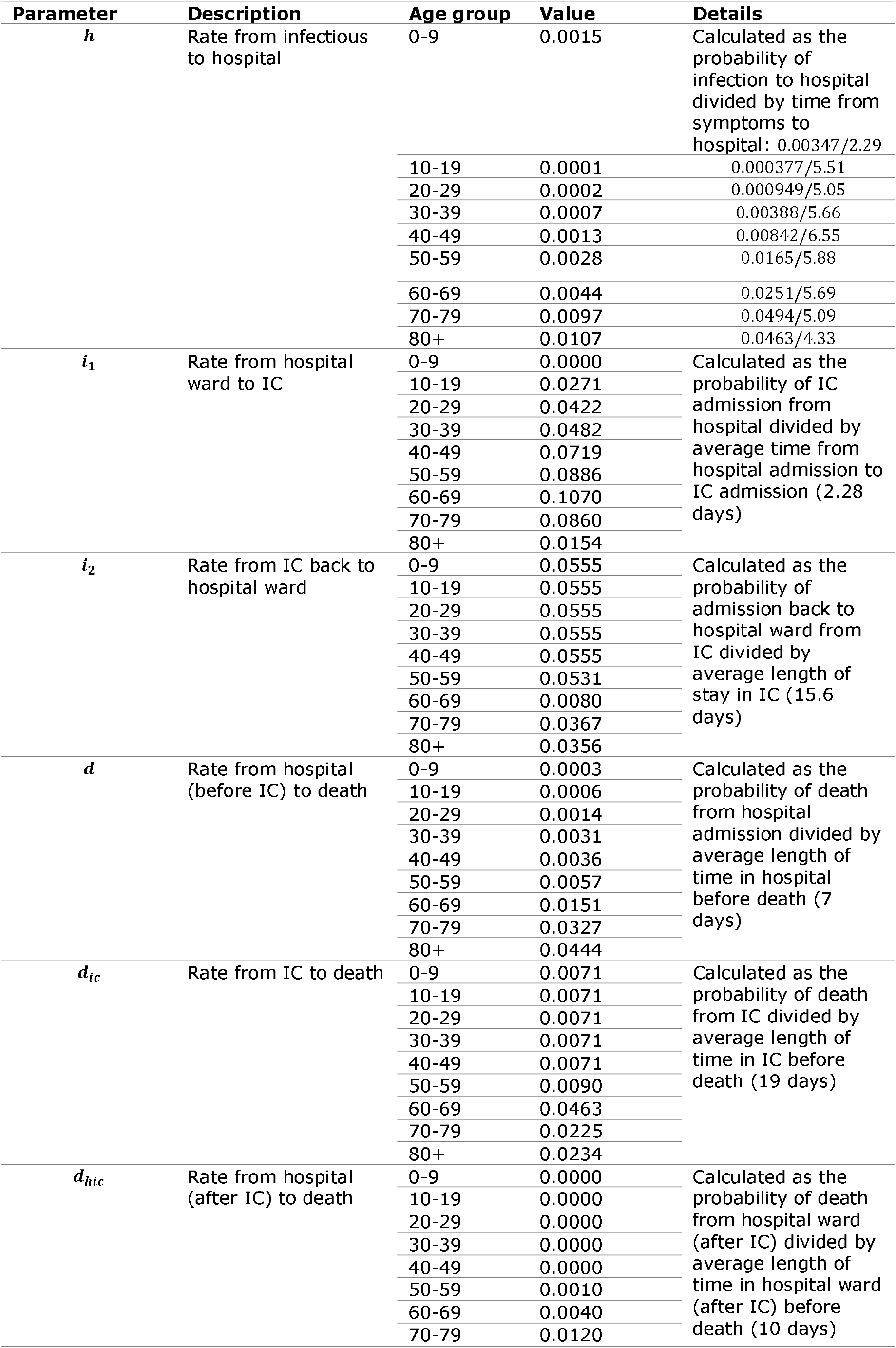
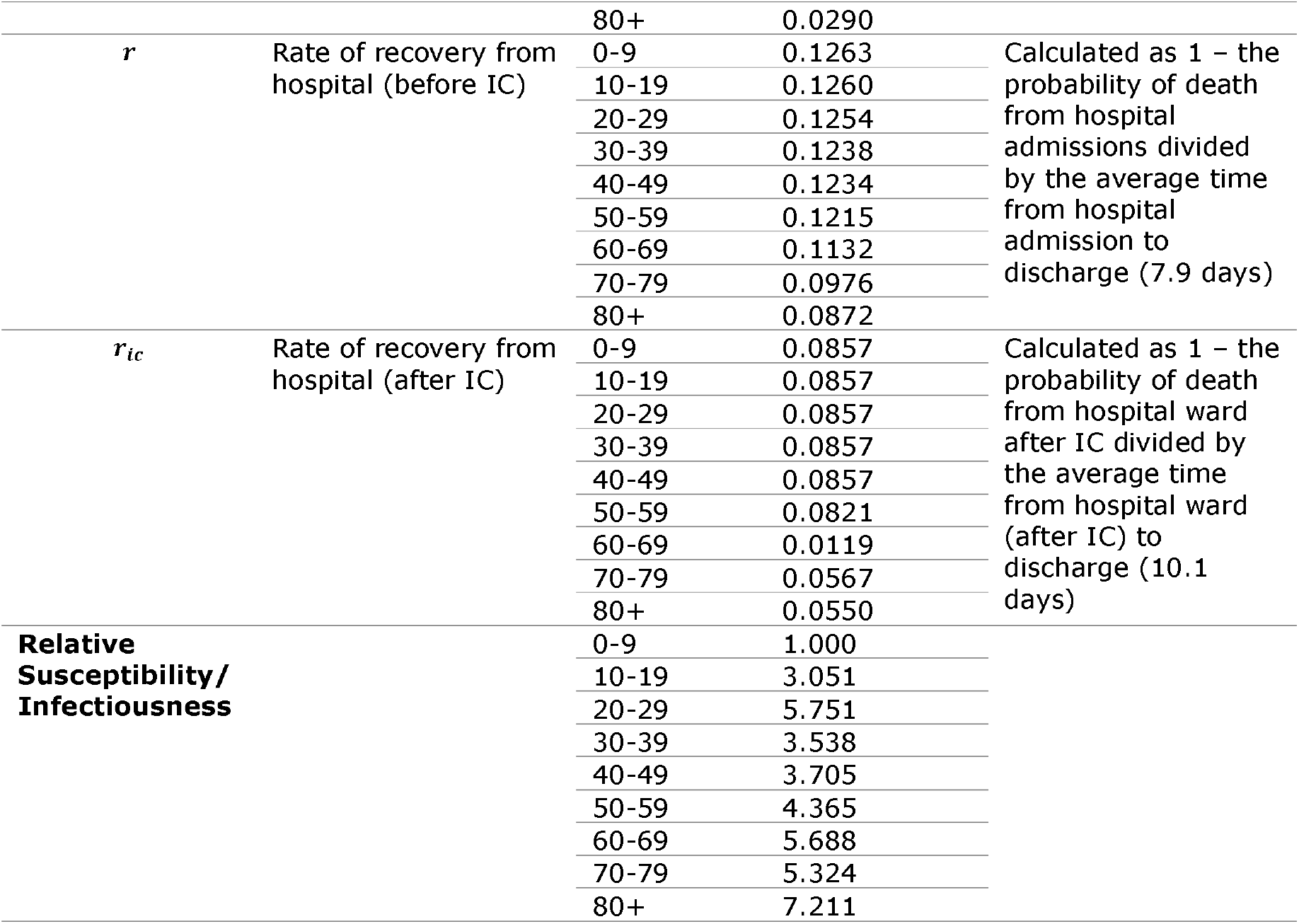
Age-dependent model parameters.

## Results

In our analysis using model parameters consistent with the Alpha variant and no waning of vaccine-induced immunity, we found reductions of 37.6% (37.6%, 37.4%) cases, a 37.5% (37.5%, 37.4%) reduction in hospital admissions and a 42.3% (42.3%, 42.1%) reduction in IC admissions in the 10–19 age group compared to no vaccination of 12-17 year olds (Figure S5, Figure S6 Table S5). This corresponded to an absolute reduction in approximately 75589 (73615, 77134) cases, 31 (30, 32) hospitalizations, and 6 (6, 6) IC admissions. Due to the reduced risk of severe outcomes in adolescents, the reduction in hospital admissions and IC admissions in the 10-19 year old group is more modest than the reduction in cases. In the remainder of the population (those aged 1-9 and >20) taken together, we observed a 7.9% (7.9%, 7.7%) reduction in cases, a 5.4% (5.4%, 5.2%) reduction in hospital admissions, and a 5.4% (5.4%, 5.2%) reduction in IC admissions under the 12+ vaccination scenario compared to the 18+ vaccination scenario (Figure S5, Figure S6, Table S5). This corresponded to an absolute reduction in 84003 (81372, 83545) cases, 1197 (1159, 1173) hospital admissions, and 319 (310, 313) IC admissions (Table S5).

To determine the robustness of our results, we performed sensitivity analyses in which we assumed vaccine-induced immunity waned completely after 1 year. Firstly, we observed a larger epidemic compared to when no waning was assumed (Figure S5, Figure S6). Secondly, adolescent vaccination resulted in more modest reductions in disease outcomes in the 10-19 year-old age group and in the remainder of the population when vaccine-induced immunity waned compared to when this immunity was stable. We observed reductions of 31.0% (31.2%, 30.9%) cases, 31.0% (31.2%, 30.9%) hospital admissions, and 37.0% (37.2%, 37.0%) IC admissions in the 10-19 year age group (Table S5). In the remainder of the population we observed reductions of 3.3% (3.6%, 3.2%) cases, 1.5% (1.8%, 1.4%) hospital admissions, and 1.9% (2.1%, 1.8%) IC admissions (Table S5).

**Table S5.**
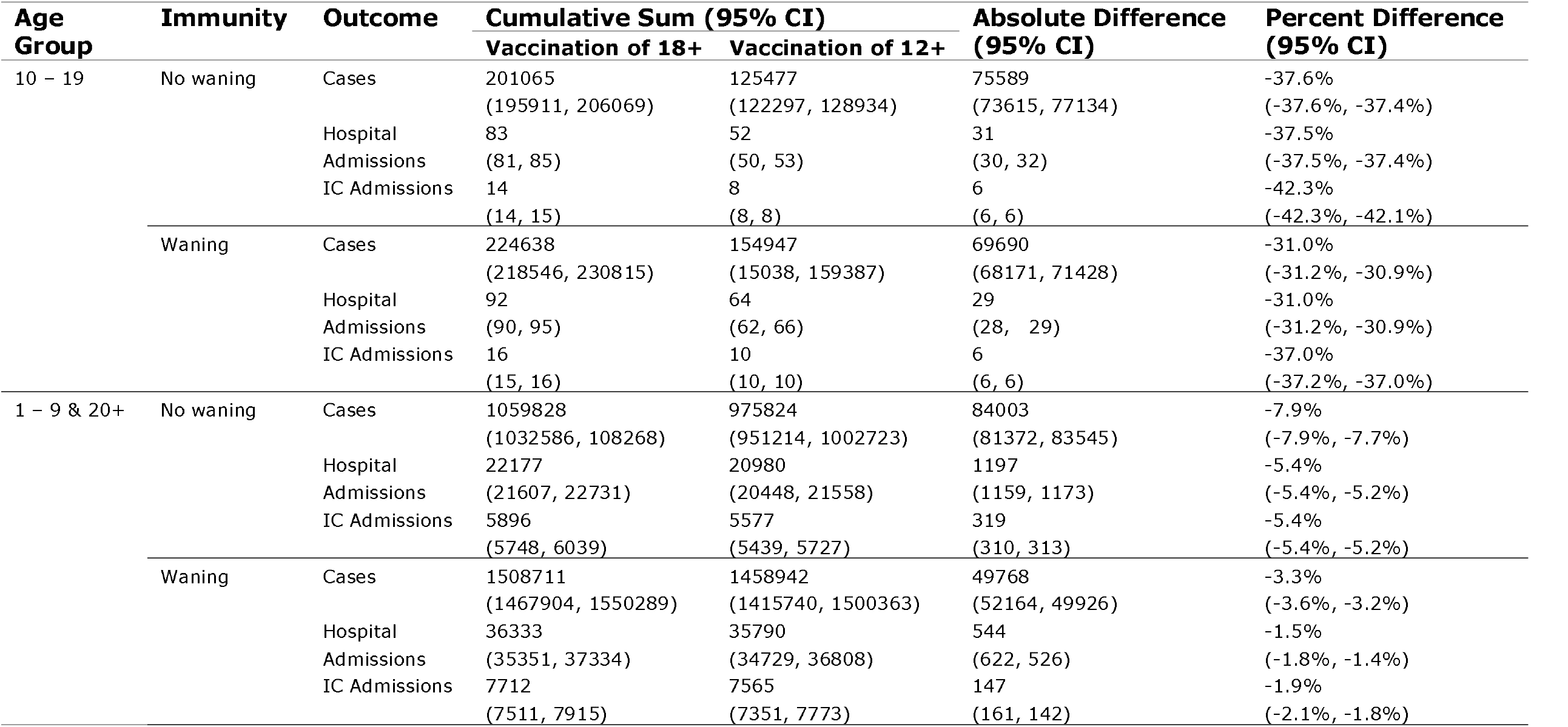
Cumulative sum, absolute difference, and percent difference of modelled outcomes with and without vaccination in 12 – 17 year olds using model parameter values consistent with the Alpha variant. Vaccination in 18+ is used as the reference for percent difference. Absolute difference is calculated as outcome in 18+ minus outcome in 12+. IC = intensive care, CI = confidence interval.

**Figure S4.**
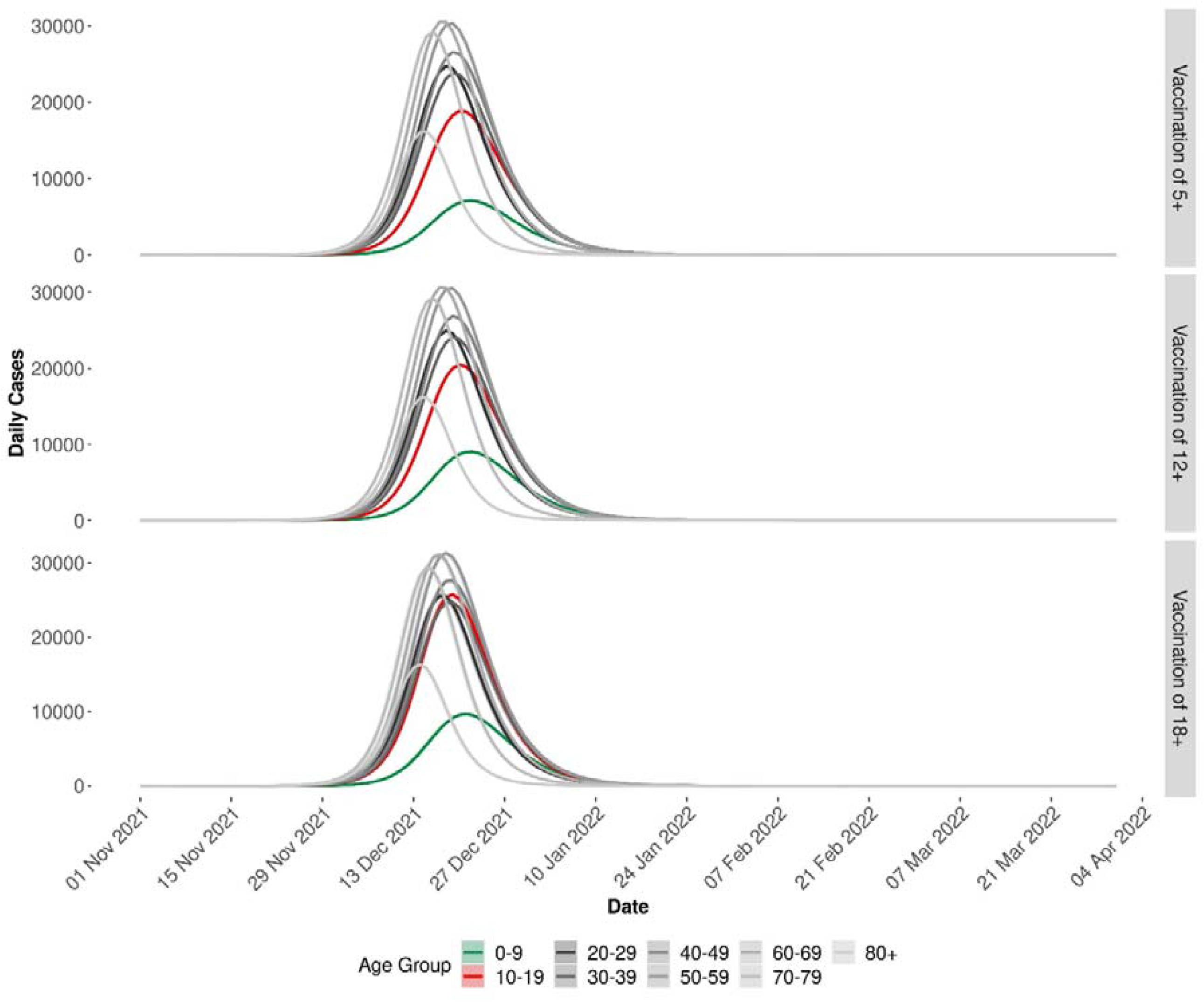
Daily cases by age group by vaccination scenario when vaccine-induced immunity is assumed to wane to zero after 1 year. Simulations were run from 22 June 2021 until 31 March 2022.

**Figure S5.**
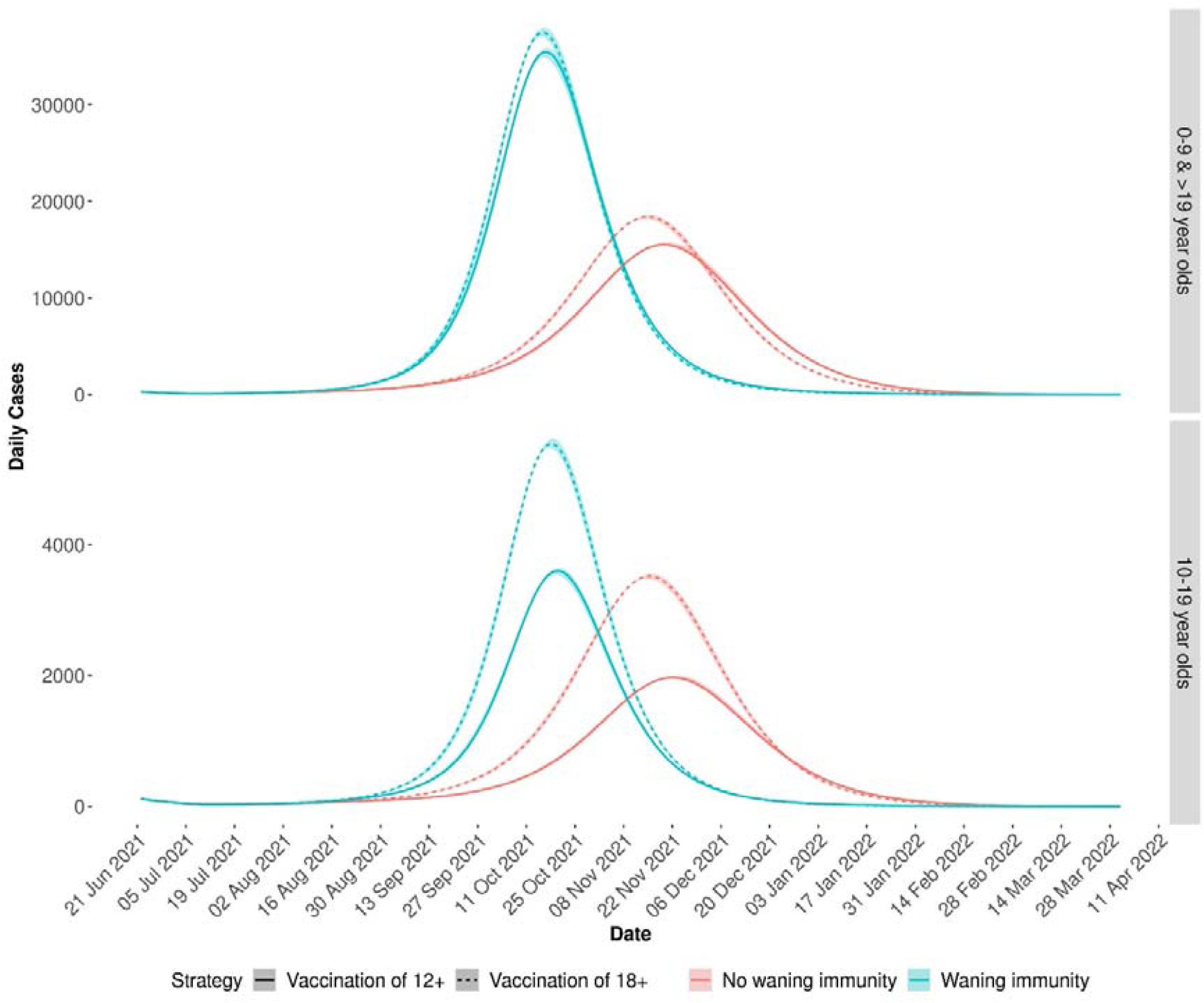
Daily cases in all individuals aged 0-9 & >19 (top) and 10-19 (bottom) when model parameters consistent with the Alpha variant were used under different vaccination scenarios: 12+ (solid line) and 18+ (dashed line). We first assumed that vaccine-induced immunity does not wane (red line) and then assumed that vaccine induced immunity waned to nothing after 1 year (blue line).

**Figure S6.**
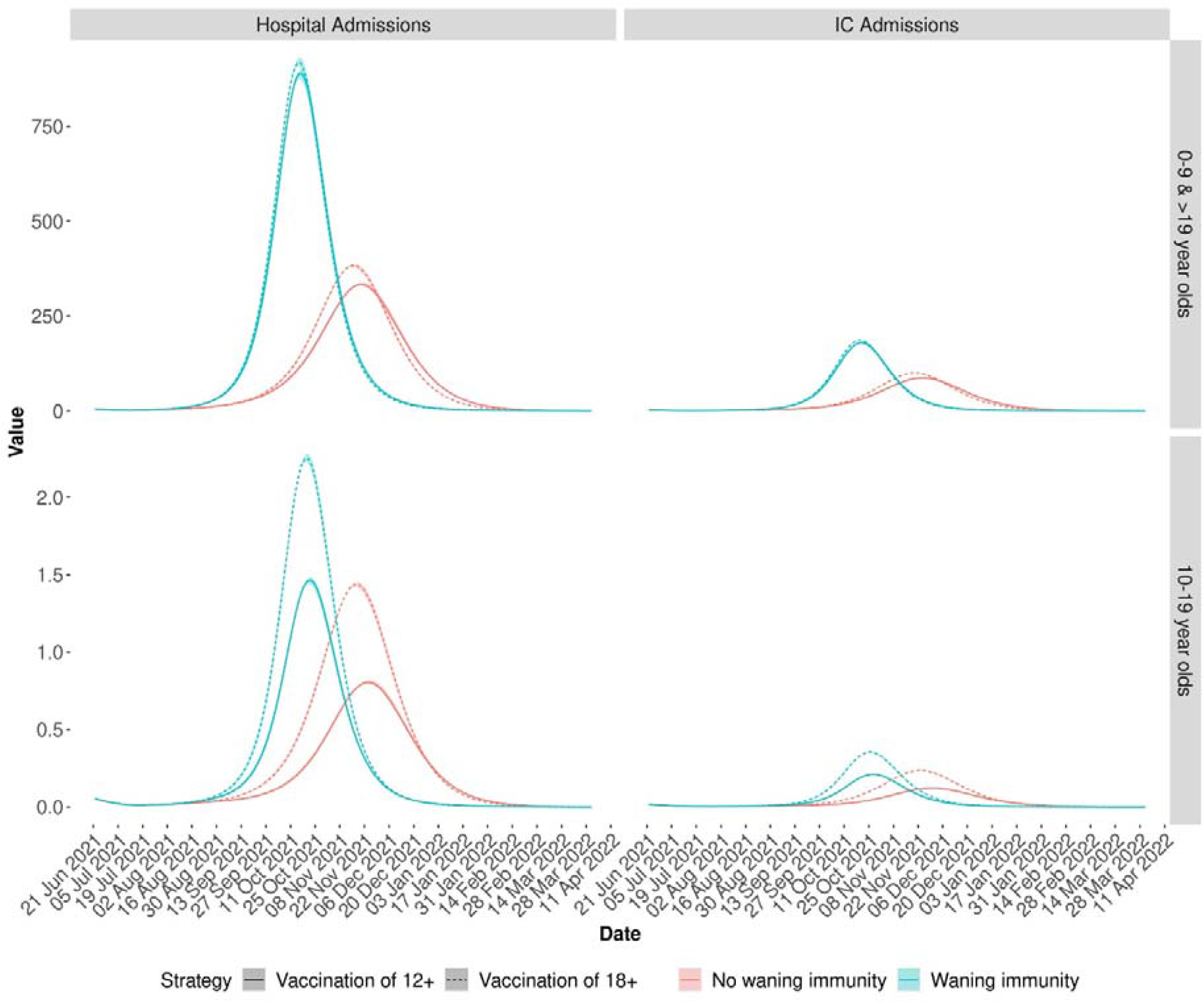
Hospital admissions and intensive care admissions in all individuals aged 0-9 & >19 (top) and 10-19 (bottom) when model parameters consistent with the Alpha variant were used under different vaccination scenarios: 12+ (solid line) and 18+ (dashed line). We first assumed that vaccine-induced immunity does not wane (red line) and then assumed that vaccine induced immunity waned to nothing after 1 year (blue line).

## Data Availability

All data and code produced are available online at: https://github.com/kylieainslie/vacamole

